# Subtype dynamics reveal horizon-dependent structure in influenza predictability

**DOI:** 10.64898/2026.05.28.26354347

**Authors:** Yicheng Mao, Benjamin Lopman, Katia Koelle, Max SY Lau

## Abstract

Accurate forecasting of seasonal influenza is critical for public health preparedness, and data-driven models are central to this effort. However, most approaches rely on aggregate indicators of influenza-like-illness (ILI), which can obscure heterogeneity and limit predictability at longer horizons. While subtype dynamics are well established, their role in data-driven forecasting remains incompletely understood. Here, we integrate subtype-resolved surveillance data into diverse data-driven frameworks using over a decade of U.S. surveillance records to evaluate and decompose predictive signal in influenza forecasting. Across pre- and post-COVID-19 periods, subtype-informed models consistently improve over baseline models trained on aggregate ILI alone, with the largest gains at longer horizons. Decomposition reveals a horizon-dependent reorganization of predictability: autoregressive persistence in recent aggregate incidence dominates at short horizons but declines with lead time, while predictive signal shifts toward subtype-derived structure. Within this structure, interaction-related features among co-circulating subtypes grow systematically with forecast horizon, indicating that longer-term predictability is driven increasingly by interaction structure rather than marginal subtype composition alone. Together, our results show that subtype information provides non-redundant predictive signal and extends the effective forecasting window of data-driven models. More broadly, our findings suggest that aggregation of heterogeneous subtype processes can obscure latent predictability, supporting subtype-resolved surveillance.

## 1 Introduction

Seasonal influenza imposes a substantial global health burden, causing recurrent waves of morbidity and healthcare strain [1, 2]. Accurate forecasting is therefore critical for public health decision-making, including timely hospital preparedness and resource allocation [3, 4]. Yet, despite substantial methodological progress in recent decades, particularly in data-driven models, forecast performance remains uneven across seasons and settings [5, 6], underscoring persistent challenges in anticipating epidemic dynamics.

These challenges reflect not only limitations in model design, but also how predictive information is structured in epidemic systems and captured in surveillance data [7]. Most data-driven approaches rely predominantly on aggregate indicators, such as influenza-like illness (ILI) rates [8, 9]. While widely available, these signals provide strong short-term autoregressive information but collapse heterogeneous transmission processes into a single population-level time series, implicitly assuming viral homogeneity and obscuring the internal structure of viral circulation [10]. As a result, the ability to resolve predictive structure may be limited, particularly at longer forecast horizons.

In practice, major surveillance platforms, such as the Centers for Disease Control and Prevention (CDC) FluView, report circulating viruses by subtype [11]. These subtypes differ in transmissibility, immune escape, and temporal dynamics [12, 13], and they co-circulate and compete for susceptible hosts through immune-mediated interactions, including partial cross-protection and short-term viral interference [14–16]. Together, these processes generate a structured, multi-component epidemic system in which interactions among subtypes can shape population-level trajectories.

However, this subtype-resolved structure has been only partially incorporated into forecasting. To date, subtype dynamics have been explicitly modeled primarily within mechanistic frameworks [17–19], which often rely on strong assumptions that limit scalability across locations and seasons. Conversely, most data-driven approaches continue to rely on aggregate incidence signals, potentially overlooking subtype-level composition and interaction that may encode additional predictive structure [20, 21]. As data-driven methods have become central tools for epidemic forecasting [22], a fundamental question remains: *does subtype-resolved surveillance provide non-redundant predictive signal beyond aggregate incidence, and how is this signal organized across forecast horizons?*

In this study, we evaluate the predictive utility of subtype-resolved surveillance by integrating virological composition into data-driven models and decomposing predictive signal into aggregate and subtype-derived components to characterize how predictability is structured across timescales. Across diverse forecasting frameworks, subtype-informed models improve forecast performance relative to models trained on aggregate incidence alone, with the clearest gains emerging at longer forecast horizons. These improvements reflect a systematic redistribution of predictive signal: the contribution of autoregressive persistence in recent aggregate incidence declines with lead time, while predictive signal increasingly shifts toward subtype-derived features, particularly interaction-related components among co-circulating subtypes. These patterns are further recapitulated in controlled simulation experiments using heterogeneous multi-subtype epidemic dynamics.

Together, our results show that subtype resolution provides non-redundant predictive information and reveal how the sources of predictability reorganize across timescales. More broadly, they provide quantitative evidence that expanding access to subtype-resolved surveillance could extend the effective forecasting horizon and improve the robustness and interpretability of data-driven epidemic forecasts.

## 2 Results

### 2.1 Temporal patterns of influenza activity and subtype structure

We first examine U.S. influenza surveillance patterns spanning the 2010–2011 through the 2024–2025 seasons to assess whether subtype-resolved structure is associated with subsequent epidemic trajectories at a descriptive level. The period from late 2018 to late 2022 was excluded because influenza circulation and surveillance were substantially disrupted during the COVID-19 pandemic. Circulating viruses were grouped into four categories, A(2009 H1N1), A(H3), A(other), and B, to characterize subtype composition over time. Figure 1 presents nationally aggregated descriptive summaries of influenza activity and subtype composition. Detailed descriptions of data sources, ILI+ construction, and preprocessing procedures are provided in *Methods: Data sources and preprocessing* .

**Fig. 1.**
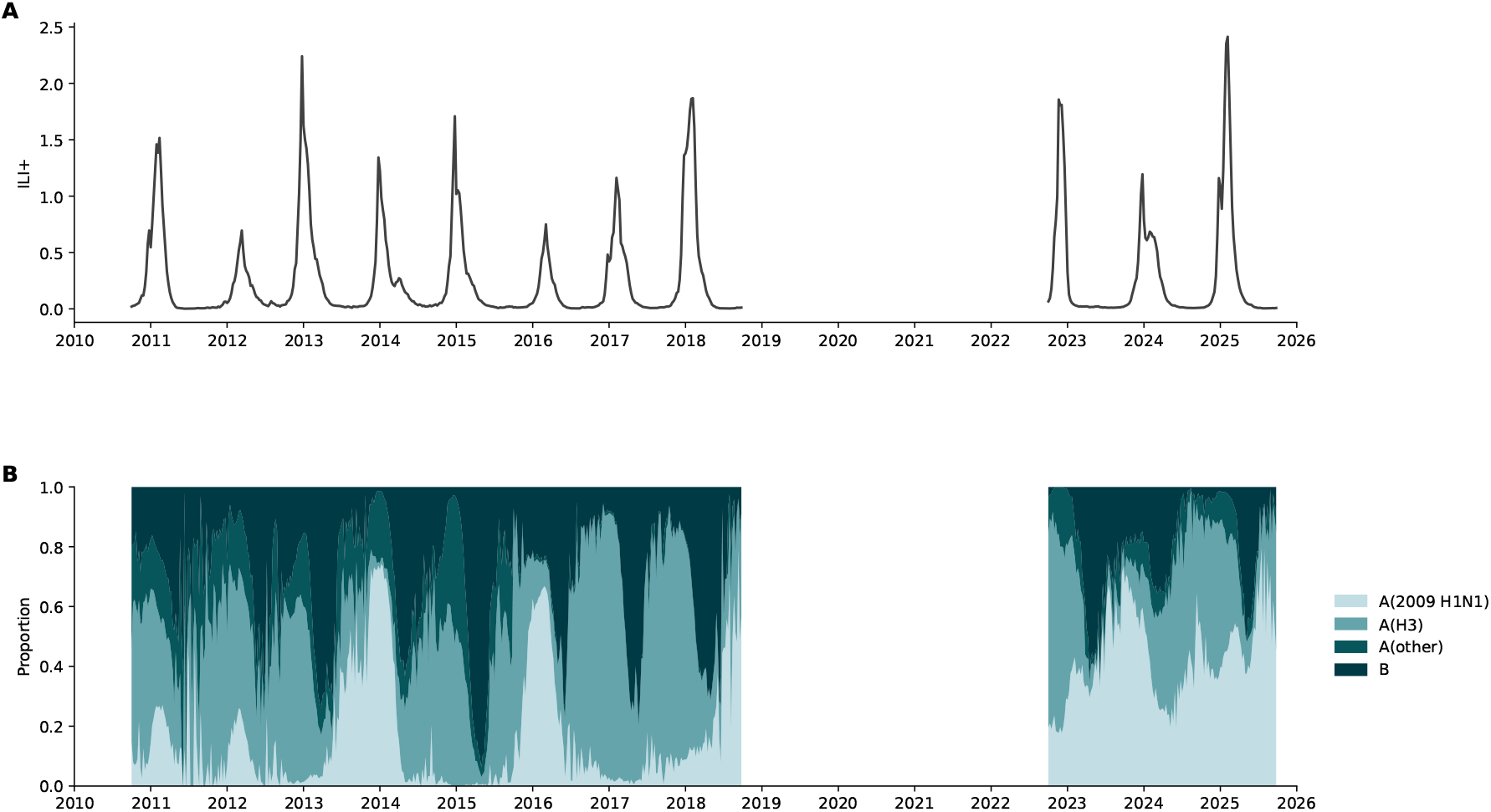
Temporal heterogeneity in seasonal influenza activity and subtype composition. **A** Weekly national ILI+ time series, where ILI+ combines ILI activity with influenza test positivity. **B** National weekly proportional circulation of influenza subtypes, where subtype proportions were normalized to sum to one within each week.

Figure 1 reveals substantial temporal heterogeneity in both epidemic intensity and subtype composition. While ILI+ exhibits clear seasonal recurrence at the national level (Fig. 1A), epidemic intensity varies markedly across years. At the compositional level, national subtype proportions display pronounced within-season restructuring (Fig. 1B). Rather than remaining static, subtype configurations evolve dynamically, with periods of relatively balanced co-circulation followed by phases in which one subtype becomes more dominant.

To further examine whether contemporaneous subtype structure is associated with subsequent epidemic trajectories, we summarized changes in ILI+ over the subsequent 1–4 weeks across observations from the 10 U.S. Department of Health and Human Services (HHS) regions (Fig. 2). For these descriptive analyses, observations were grouped by dominant subtype state and by overall interaction intensity, as defined in *Methods: Descriptive summaries of subtype structure*. As shown in Fig. 2A, observations in which A(H3) was the dominant subtype were, on average, followed by larger increases in ILI+ over the subsequent 1–4 weeks, whereas the remaining dominant-state groups generally showed flatter or declining trajectories. A similar pattern is observed when grouping observations by overall subtype interaction intensity (Fig. 2B): observations with lower interaction tend to be followed by increasing ILI+, whereas those with higher interaction are more often associated with stable or declining trajectories. The magnitude and direction of these associations vary across forecast horizons, suggesting that the relationship between subtype structure and subsequent trajectories may depend on prediction timescale.

**Fig. 2.**
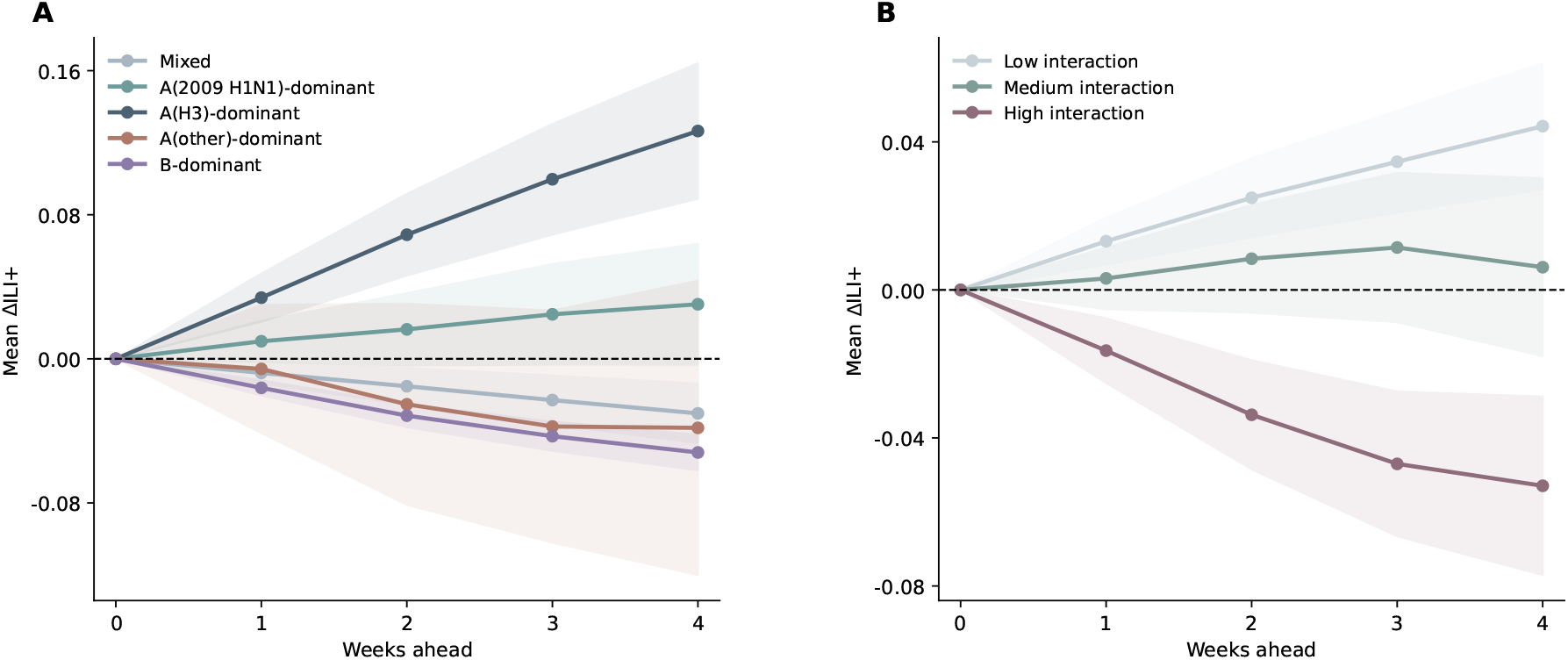
Strong associations of contemporaneous subtype structure with subsequent trajectories of aggregated ILI+ at the HHS regional level. **A** Mean change in aggregated ILI+ over the subsequent 1–4 weeks, stratified by contemporaneous dominant subtype state. **B** Mean change in aggregated ILI+ over the subsequent 1–4 weeks, stratified by contemporaneous subtype interaction intensity. Here, ΔILI+ denotes the change in aggregated ILI+ relative to week *t*. Shaded bands indicate 95% confidence intervals around the group means. Definitions of dominant subtype state, interaction intensity, and trajectory summaries are provided in Methods: Descriptive summaries of subtype structure.

Taken together, these descriptive patterns suggest that subtype-resolved information may provide additional predictive signal beyond contemporaneous composition alone. This provides descriptive motivation for incorporating subtype-resolved signals and interaction structure into data-driven models, rather than collapsing influenza activity into a single aggregate incidence series.

### 2.2 Horizon-dependent contribution of subtype information to predictability

We first evaluated whether subtype-resolved surveillance signals improve forecasting of weekly ILI+ incidence at the HHS regional level relative to baseline autoregressive models trained solely on aggregate ILI+, and whether their contribution varies across forecast horizons. Baseline models used only the most recent aggregate ILI+ intensity as the predictor, whereas subtype-augmented models extended the predictor set by integrating subtype proportions as additional features. Forecasting was performed using two tree-based learners (Random Forest [23] and XGBoost [24]), two recurrent neural networks (LSTM [25] and GRU [26]), and an unweighted ensemble combining their predictions. Forecast performance was evaluated using root mean squared error (RMSE), Pearson correlation, peak week error and weighted interval score (WIS).

Together, these metrics capture complementary aspects of predictive quality, including magnitude accuracy, temporal concordance between predicted and observed trajectories, peak timing alignment, and probabilistic uncertainty assessment. Detailed descriptions of feature construction, model specification, training procedures, and evaluation metrics are provided in the *Methods* section under *Feature engineering, Model frameworks and training*, and *Evaluation metrics*.

Table 1 summarizes forecasting performance across 1–4 week ahead horizons for both pre-COVID and post-COVID periods. All metrics were computed separately for each of the 10 HHS regions and then averaged across regions. Across both periods, absolute predictive accuracy declined monotonically with increasing forecast horizon, reflecting the increasing difficulty of forecasting influenza at longer lead times. This horizon effect was consistent across model classes and evaluation metrics, and is in line with prior influenza forecasting evaluations showing that predictive accuracy generally deteriorates as lead time extends [9].

**Table 1.** Forecasting performance at 1–4 week ahead horizons. Results are shown separately for the pre-COVID and post-COVID periods. Metrics were computed separately for each of the 10 HHS regions and then averaged across regions. For RMSE and WIS, 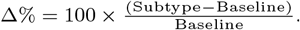. For Pearson Correlation and Peak Error, Δ = Subtype − Baseline. Negative Δ% indicates reduced error (RMSE/WIS) under the subtype model. Positive Δ in Pearson Correlation indicates improved correlation. Negative Δ in Peak Error indicates reduced peak timing error. Bold Δ entries indicate improvement of the subtype model over the baseline for the ensemble.

| Horizon | Model | RMSE |  |  | Pearson Correlation |  |  | Peak Error (weeks) |  |  | WIS |  |  |
| --- | --- | --- | --- | --- | --- | --- | --- | --- | --- | --- | --- | --- | --- |
| | | Baseline | Subtype | $\Delta\%$ | Baseline | Subtype | $\Delta$ | Baseline | Subtype | $\Delta$ | Baseline | Subtype | $\Delta\%$ |
| **Before COVID-19** |  |  |  |  |  |  |  |  |  |  |  |  |  |
| 1W | RF | 0.220 | 0.244 | 11.11 | 0.948 | 0.956 | 0.008 | 1.3 | 1.5 | 0.2 | 0.090 | 0.100 | 10.43 |
| 1W | XGB | 0.231 | 0.224 | -3.12 | 0.942 | 0.953 | 0.011 | 1.2 | 1.4 | 0.2 | 0.097 | 0.096 | -1.65 |
| 1W | LSTM | 0.205 | 0.188 | -8.45 | 0.949 | 0.958 | 0.009 | 1.0 | 1.1 | 0.1 | 0.085 | 0.076 | -10.10 |
| 1W | GRU | 0.212 | 0.190 | -10.10 | 0.949 | 0.958 | 0.009 | 1.0 | 1.4 | 0.4 | 0.087 | 0.077 | -11.50 |
| 1W | **Ensemble** | 0.214 | 0.208 | **-2.96** | 0.949 | 0.958 | **0.009** | 1.0 | 1.3 | 0.3 | 0.089 | 0.086 | **-3.35** |
| 2W | RF | 0.354 | 0.349 | -1.38 | 0.841 | 0.882 | 0.041 | 2.1 | 2.0 | -0.1 | 0.147 | 0.143 | -3.06 |
| 2W | XGB | 0.355 | 0.348 | -1.89 | 0.845 | 0.868 | 0.022 | 1.6 | 2.0 | 0.4 | 0.151 | 0.147 | -2.64 |
| 2W | LSTM | 0.368 | 0.338 | -8.28 | 0.848 | 0.891 | 0.043 | 2.0 | 1.5 | -0.5 | 0.153 | 0.136 | -11.22 |
| 2W | GRU | 0.363 | 0.345 | -5.17 | 0.850 | 0.892 | 0.042 | 2.0 | 1.5 | -0.5 | 0.152 | 0.139 | -8.31 |
| 2W | **Ensemble** | 0.357 | 0.342 | **-4.34** | 0.847 | 0.885 | **0.038** | 2.0 | 1.8 | **-0.2** | 0.150 | 0.140 | **-6.37** |
| 3W | RF | 0.447 | 0.416 | -6.91 | 0.736 | 0.836 | 0.099 | 3.0 | 2.3 | -0.7 | 0.190 | 0.172 | -9.54 |
| 3W | XGB | 0.448 | 0.423 | -5.67 | 0.745 | 0.814 | 0.069 | 2.2 | 2.3 | 0.1 | 0.196 | 0.179 | -8.65 |
| 3W | LSTM | 0.460 | 0.437 | -4.93 | 0.745 | 0.854 | 0.109 | 3.1 | 1.8 | -1.3 | 0.195 | 0.179 | -8.54 |
| 3W | GRU | 0.459 | 0.444 | -3.35 | 0.745 | 0.861 | 0.116 | 3.1 | 1.8 | -1.3 | 0.195 | 0.181 | -6.95 |
| 3W | **Ensemble** | 0.452 | 0.426 | **-5.82** | 0.744 | 0.849 | **0.104** | 2.7 | 1.7 | **-1.0** | 0.193 | 0.175 | **-9.19** |
| 4W | RF | 0.507 | 0.458 | -9.50 | 0.624 | 0.812 | 0.188 | 3.7 | 1.7 | -2.0 | 0.221 | 0.191 | -13.60 |
| 4W | XGB | 0.512 | 0.471 | -7.93 | 0.648 | 0.800 | 0.152 | 2.6 | 1.8 | -0.8 | 0.227 | 0.199 | -12.47 |
| 4W | LSTM | 0.523 | 0.507 | -3.16 | 0.646 | 0.836 | 0.190 | 4.5 | 1.5 | -3.0 | 0.226 | 0.210 | -7.11 |
| 4W | GRU | 0.522 | 0.516 | -1.25 | 0.645 | 0.823 | 0.178 | 4.5 | 1.7 | -2.8 | 0.226 | 0.214 | -5.37 |
| 4W | **Ensemble** | 0.513 | 0.482 | **-6.09** | 0.645 | 0.842 | **0.198** | 2.9 | 1.5 | **-1.4** | 0.223 | 0.200 | **-10.50** |
| **After COVID-19** |  |  |  |  |  |  |  |  |  |  |  |  |  |
| 1W | RF | 0.239 | 0.239 | 0.13 | 0.930 | 0.937 | 0.007 | 1.4 | 1.3 | -0.1 | 0.096 | 0.095 | -1.57 |
| 1W | XGB | 0.234 | 0.232 | -0.48 | 0.934 | 0.939 | 0.006 | 0.9 | 1.0 | 0.1 | 0.096 | 0.096 | -0.85 |
| 1W | LSTM | 0.232 | 0.230 | -0.94 | 0.939 | 0.938 | -0.002 | 1.0 | 1.0 | 0.0 | 0.093 | 0.092 | -1.32 |
| 1W | GRU | 0.230 | 0.226 | -1.79 | 0.939 | 0.937 | -0.001 | 1.0 | 1.0 | 0.0 | 0.092 | 0.092 | -0.31 |
| 1W | **Ensemble** | 0.231 | 0.228 | **-1.25** | 0.937 | 0.939 | **0.003** | 1.0 | 1.0 | 0.0 | 0.093 | 0.092 | **-1.52** |
| 2W | RF | 0.384 | 0.370 | -3.52 | 0.809 | 0.840 | 0.030 | 2.5 | 2.3 | -0.2 | 0.157 | 0.148 | -5.75 |
| 2W | XGB | 0.377 | 0.384 | 1.78 | 0.817 | 0.812 | -0.005 | 1.7 | 2.8 | 1.1 | 0.157 | 0.155 | -1.41 |
| 2W | LSTM | 0.377 | 0.364 | -3.32 | 0.827 | 0.828 | 0.001 | 2.0 | 2.0 | 0.0 | 0.153 | 0.150 | -1.82 |
| 2W | GRU | 0.376 | 0.364 | -3.30 | 0.826 | 0.829 | 0.003 | 2.0 | 2.0 | 0.0 | 0.153 | 0.150 | -1.51 |
| 2W | **Ensemble** | 0.376 | 0.364 | **-3.21** | 0.821 | 0.831 | **0.010** | 2.1 | 2.0 | **-0.1** | 0.154 | 0.148 | **-3.65** |
| 3W | RF | 0.467 | 0.457 | -2.17 | 0.708 | 0.738 | 0.030 | 3.4 | 3.2 | -0.2 | 0.198 | 0.189 | -4.36 |
| 3W | XGB | 0.463 | 0.455 | -1.77 | 0.712 | 0.739 | 0.027 | 2.7 | 3.4 | 0.7 | 0.198 | 0.191 | -3.53 |
| 3W | LSTM | 0.463 | 0.443 | -4.26 | 0.731 | 0.736 | 0.005 | 2.8 | 3.0 | 0.2 | 0.193 | 0.187 | -2.90 |
| 3W | GRU | 0.462 | 0.440 | -4.85 | 0.730 | 0.744 | 0.014 | 3.1 | 3.0 | -0.1 | 0.193 | 0.186 | -3.54 |
| 3W | **Ensemble** | 0.461 | 0.442 | **-4.12** | 0.723 | 0.747 | **0.025** | 3.5 | 3.0 | **-0.5** | 0.194 | 0.185 | **-4.56** |
| 4W | RF | 0.518 | 0.500 | -3.45 | 0.633 | 0.701 | 0.068 | 4.1 | 3.6 | -0.5 | 0.227 | 0.211 | -7.35 |
| 4W | XGB | 0.518 | 0.534 | 3.14 | 0.630 | 0.614 | -0.016 | 3.5 | 4.2 | 0.7 | 0.229 | 0.224 | -2.04 |
| 4W | LSTM | 0.520 | 0.488 | -6.10 | 0.660 | 0.684 | 0.023 | 4.3 | 3.3 | -1.0 | 0.222 | 0.212 | -4.47 |
| 4W | GRU | 0.518 | 0.499 | -3.81 | 0.658 | 0.654 | -0.004 | 4.0 | 3.9 | -0.1 | 0.222 | 0.222 | -0.09 |
| 4W | **Ensemble** | 0.516 | 0.494 | **-4.14** | 0.649 | 0.690 | **0.041** | 4.1 | 3.3 | **-0.8** | 0.223 | 0.212 | **-4.91** |

At the 1-week horizon prior to COVID-19, the benefit of subtype information was limited and architecture dependent. Compared with the corresponding models trained on aggregated ILI+ alone, neural models (LSTM and GRU) achieved moderate reductions in RMSE and WIS of approximately 8–11%, whereas tree-based learners (RF and XGB) showed smaller or inconsistent changes. Gains in Pearson correlation were modest, and peak timing error remained largely unchanged.

In contrast, a more systematic subtype effect emerged at the 3–4-week horizons prior to COVID-19. At 3–4 weeks ahead, subtype-augmented models consistently reduced both RMSE and WIS across most architectures, with percentage reductions typically ranging from 5–14% depending on model class. The ensemble achieved a 6.09% RMSE reduction and a 10.50% WIS reduction at 4 weeks. Pearson correlations increased substantially at these horizons, with gains exceeding 0.10 for several models and reaching 0.198 for the ensemble at 4 weeks. Improvements were also evident in peak timing error, with subtype models showing 1–3 fewer weeks of error in the predicted peak week for several architectures. These results indicate that subtype resolution enhanced overall trajectory fidelity, probabilistic sharpness and calibration, and the anticipation of epidemic turning points.

After COVID-19, overall predictability declined and performance differences narrowed. Short-horizon improvements remained limited across RMSE, WIS, correlation, and peak timing metrics. However, subtype-augmented models continued to yield consistent gains at 3–4 weeks ahead. At the 4-week horizon, the ensemble reduced RMSE by 4.14%, reduced WIS by 4.91%, increased Pearson correlation by 0.041, and reduced peak timing error by 0.8 weeks relative to the baseline. Although attenuated compared to the pre-COVID period, these improvements demonstrate that subtype information retained predictive value at longer horizons under altered (post-COVID) transmission dynamics.

To complement the region-averaged summaries above, we provide region-specific patterns in the *Supplementary Information*. These analyses support the overall conclusions, while highlighting some heterogeneity: subtype information may, in certain settings, appear detrimental for specific model classes, particularly tree-based methods, in short-term predictions for some HHS regions.

Taken together, these results show that subtype resolution primarily enhances forecasting at longer horizons, whereas its contribution is more limited at short lead times.

#### Robustness analyses: extended forecast horizons and delayed subtype information

To assess the robustness of this horizon-dependent advantage, we conducted two additional analyses. First, we extended the forecast horizon to 5–8 weeks ahead. Although absolute predictive accuracy declined at longer lead times, subtype-augmented models consistently outperformed baseline models across most architectures. Second, recognizing that subtype reports may be available with delay in operational settings, we constructed subtype features using proportions lagged by an additional week relative to ILI+. Even under this delayed-information scenario, the relative advantage of subtype-augmented models persisted across RMSE, correlation, and peak timing metrics. Detailed quantitative results are provided in the *Supplementary Information*.

### 2.3 Decomposition of predictive signal reveals horizon-dependent structure

To identify the sources of horizon-dependent predictability, we decompose predictive importance into three components using permutation importance [23]: autoregressive features derived from lagged (aggregate) ILI+ intensity, subtype main-effect features capturing intensity-weighted marginal subtype prevalence, and subtype interaction features capturing pairwise co-circulation structure among subtypes. Detailed computational procedures are provided in *Methods: Feature importance analysis*.

Figure 3A summarizes the share of total permutation importance attributable to each predictor group under the ensemble model across one-to four-week forecast horizons, shown separately for the pre-COVID and post-COVID periods. In the pre-COVID period, autoregressive features account for approximately 38% of total importance at the one-week horizon, declining steadily to approximately 11% at four weeks. Conversely, the combined contribution of subtype-derived predictors rises from roughly 62% to nearly 89% over the same range. A similar pattern holds in the post-COVID period, although autoregressive features retain a somewhat larger share at short horizons and the redistribution is slightly less pronounced. Across both periods, these results confirm that near-term forecasts rely substantially on recent incidence persistence, whereas longer-horizon predictions depend increasingly on subtype-resolved virological composition.

**Fig. 3.**
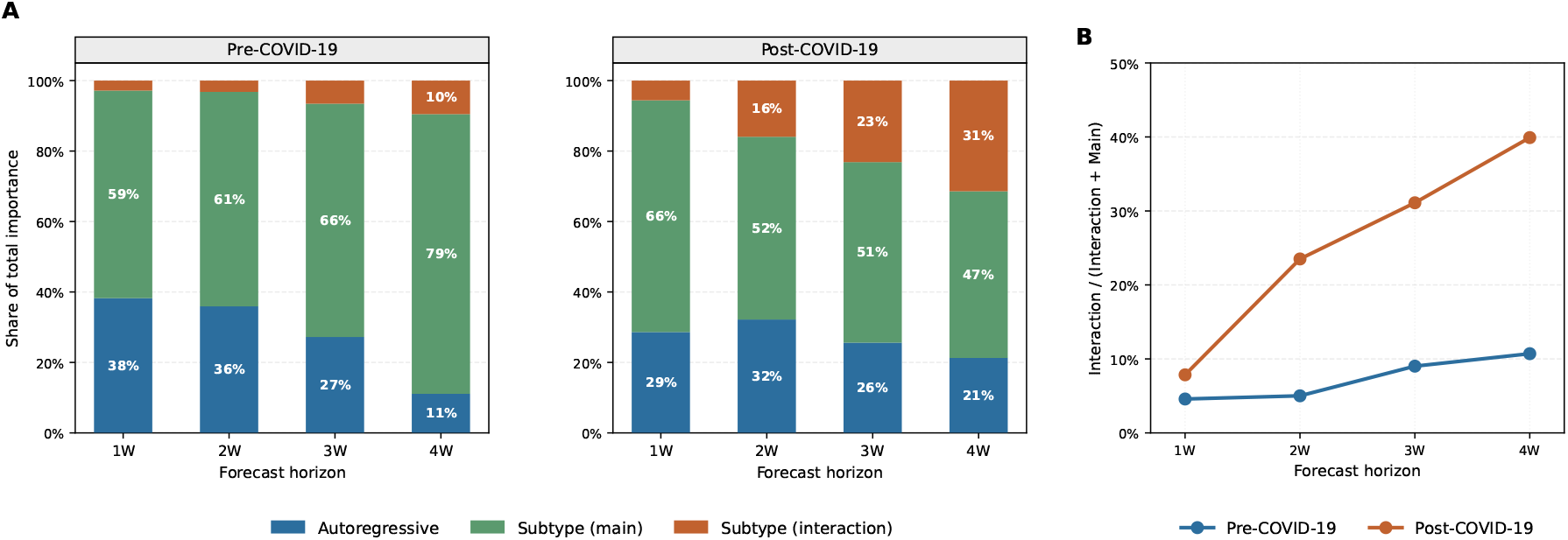
Decomposition of predictive signal across forecast horizons under the ensemble model. **A** Share of total permutation importance attributable to autoregressive features, subtype main-effect features, and subtype interaction features across one-to four-week horizons, shown separately for the pre-COVID-19 (left) and post-COVID-19 (right) periods. Importance shares are normalized to sum to one within each horizon. **B** Ratio of interaction importance to total subtype importance (interaction plus main) across forecast horizons. Higher values indicate that the growing contribution of subtype-derived information is increasingly associated with co-circulation and interaction structure rather than marginal subtype prevalence.

Importantly, the growing importance of subtype-derived information at longer horizons is not distributed uniformly across subtype predictors, but becomes increasingly concentrated in interaction-related structure. Figure 3B quantifies this internal redistribution by plotting the ratio of interaction importance to total subtype importance for the ensemble model. In the pre-COVID period, this ratio increases modestly from approximately 4% to 10% between the one- and four-week horizons. The increase is substantially more pronounced in the post-COVID period, where the interaction share rises from approximately 8% at one week to 40% at four weeks. In both periods, the ratio increases monotonically with forecast horizon, indicating that as subtype-derived predictors become more important overall, an increasing fraction of that predictive contribution is attributable to interaction structure. These results suggest that the horizon-dependent gain in predictive signal is increasingly driven by co-circulation and interaction structure among subtypes, rather than by marginal subtype composition alone.

Taken together, these findings provide a mechanistic interpretation of the forecasting gains reported in Section 2.2. The improved performance of subtype-informed models at longer horizons reflects two concurrent shifts in predictive signal: a redistribution from autoregressive to subtype-derived information, and, within the subtype component, a progressive reweighting toward interaction-driven structure. This horizon-dependent transition indicates that longer-term epidemic predictability is increasingly shaped by the competitive and interferential dynamics among co-circulating subtypes, rather than by persistence in recent aggregate incidence or by any single subtype alone.

### 2.4 Simulation study

To further assess whether subtype-driven structure can generate the horizon-dependent predictive patterns observed in the empirical analysis, we conducted a controlled simulation study based on multi-subtype influenza epidemics. Our goal is to construct a minimal multi-component epidemic system with heterogeneous dynamics, designed to test whether structured co-circulation, without explicitly modeling biological interactions, can generate the observed horizon-dependent predictability patterns.

Rather than attempting to reproduce the full biological dynamics of specific influenza subtypes, we simulated three co-circulating subtype components, denoted as subtype 1, subtype 2, and subtype 3. Parameter values for the three subtype components were selected from, or calibrated to match, published influenza estimates, so that subtype 1, subtype 2, and subtype 3 broadly approximate plausible A(H1N1)-, A(H3N2)-, and B-like seasonal influenza profiles, respectively. These literature-informed choices covered latent and infectious periods, reproduction-number ranges, seasonal forcing, epidemic timing, and susceptibility levels [12, 13, 27–32]. The model incorporates heterogeneity in subtype-specific epidemiological parameters. Detailed epidemiological specifications and implementation are provided in the *Supplementary Information*.

The simulated outbreaks exhibit structured variation in subtype co-circulation across regions and seasons (Figure 4A–B), arising from heterogeneity in subtype dynamics. Specifically, the simulations generated recurrent epidemic waves in aggregate incidence and substantial temporal variation in subtype composition. Different subtypes dominated at different times within and across seasons, reflecting heterogeneity in transmissibility, timing, and seasonality. These asynchronous subtype trajectories produce population-level epidemic patterns that resemble interaction or competition among subtypes.

**Fig. 4.**
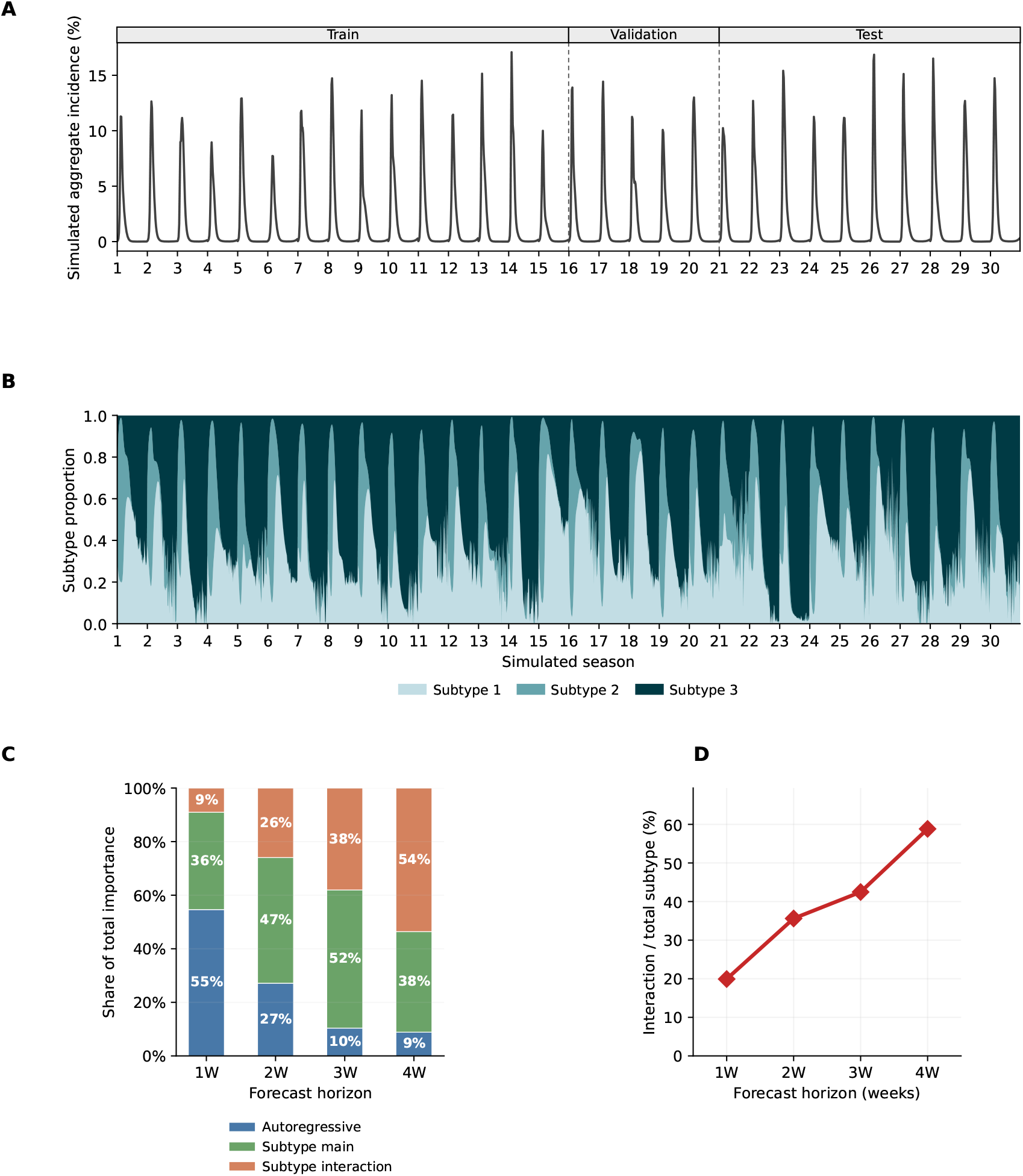
Simulation study based on stylized multi-subtype influenza epidemics. **A** Simulated aggregate weekly incidence, expressed as a percentage of the population and averaged across five regions over 30 seasons. The top strip and vertical dashed lines indicate the train, validation, and test periods used in the simulation forecasting analysis. **B** Simulated subtype composition over time, shown as the proportional contribution of subtype 1, subtype 2, and subtype 3. **C** Share of total permutation importance attributable to autoregressive, subtype main-effect, and subtype-interaction features under the ensemble model across 1–4 week forecast horizons. **D** Ratio of interaction importance to total subtype-derived importance for the ensemble model, defined as interaction importance divided by the sum of subtype main-effect and interaction importance.

Notably, we do not impose an explicit cross-immunity or competition structure, as the precise nature of subtype interactions remains incompletely understood and is not well parameterized in the literature. Instead, potential interaction structure is represented implicitly through heterogeneity in subtype-specific epidemiological dynamics, together with compositional constraints on subtype prevalence. This provides a conservative setting for evaluating whether subtype-informed predictors, including both marginal subtype composition and pairwise co-circulation terms, can recover forecast-relevant structure arising from heterogeneous subtype dynamics.

We then applied the same forecasting framework used in the preceding empirical analysis to the simulated data. Baseline models used lagged aggregate incidence only, whereas subtype-augmented models additionally included subtype main-effect and pairwise subtype-interaction features. Table 2 summarizes ensemble forecasting performance across 1–4 week forecast horizons in the simulation study. The subtype-augmented ensemble consistently improved over the baseline ensemble across all four evaluation metrics. The RMSE reduction increased from 8.18% at the 1-week horizon to 26.39% at the 4-week horizon, while the WIS reduction increased from 9.13% to 37.99%. Pearson correlation gains also increased from 0.018 to 0.338, and peak timing error decreased more substantially at longer horizons, from a reduction of 0.06 weeks at the 1-week horizon to 2.98 weeks at the 4-week horizon. These results are consistent with the empirical analysis and further support the conclusion that subtype-derived information becomes increasingly informative as forecasts move beyond short-term autoregressive persistence.

**Table 2.**
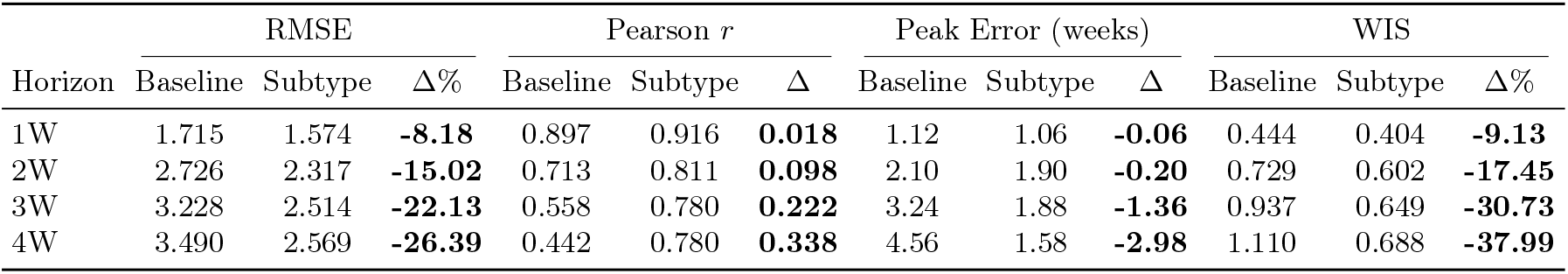
Ensemble forecasting performance in the simulation study across 1–4 week forecast horizons. Bold Δ entries indicate improvement of the subtype-augmented ensemble over the baseline ensemble.

| Horizon | RMSE | | | Pearson $r$ | | | Peak Error (weeks) | | | WIS | | |
| --- | --- | --- | --- | --- | --- | --- | --- | --- | --- | --- | --- | --- |
| | Baseline | Subtype | $\Delta\%$ | Baseline | Subtype | $\Delta$ | Baseline | Subtype | $\Delta$ | Baseline | Subtype | $\Delta\%$ |
| 1W | 1.715 | 1.574 | **-8.18** | 0.897 | 0.916 | **0.018** | 1.12 | 1.06 | **-0.06** | 0.444 | 0.404 | **-9.13** |
| 2W | 2.726 | 2.317 | **-15.02** | 0.713 | 0.811 | **0.098** | 2.10 | 1.90 | **-0.20** | 0.729 | 0.602 | **-17.45** |
| 3W | 3.228 | 2.514 | **-22.13** | 0.558 | 0.780 | **0.222** | 3.24 | 1.88 | **-1.36** | 0.937 | 0.649 | **-30.73** |
| 4W | 3.490 | 2.569 | **-26.39** | 0.442 | 0.780 | **0.338** | 4.56 | 1.58 | **-2.98** | 1.110 | 0.688 | **-37.99** |

Finally, we decomposed the predictive signal in the simulated forecasts using the same permutation-importance framework as in the preceding empirical analysis. Figure 4C–D shows a clear horizon-dependent redistribution of predictive importance under the ensemble model. Autoregressive importance declined from 55% at the 1-week horizon to 9% at the 4-week horizon, whereas subtype-interaction importance increased from 9% to 54%. The ratio of interaction importance to total subtype-derived importance also increased with forecast horizon, indicating that the subtype-derived signal became increasingly interaction-dominated at longer horizons.

These patterns are more pronounced in the simulation setting, where infection incidence and subtype composition are observed under a controlled data-generating process. This contrast suggests that the more modest and heterogeneous gains observed in the empirical analysis may partly reflect limitations of surveillance data, including observational noise, reporting variability, and the presence of less biologically resolved categories (e.g., A(other)), which can dilute subtype-derived predictive signal.

These results provide additional support for the mechanistic interpretation that longer-horizon influenza forecasts draw increasingly on subtype-informed epidemic structure, particularly structured co-circulation among subtypes, rather than on recent aggregate incidence alone.

## 3 Discussion

Data-driven models have become central tools for epidemic response and preparedness, including influenza forecasting [6]. Models based on aggregate ILI+ benefit from strong autoregressive structure, yet this dependence can constrain predictive capacity at longer horizons. As the influence of recent incidence diminishes and uncertainty accumulates, influenza forecasts become increasingly difficult [9]. Under these conditions, subtype-resolved information may provide additional predictive structure by capturing heterogeneity in transmission dynamics, compositional variation, and competitive interactions among co-circulating subtypes [17, 33]. Yet, how predictive information is structured within subtype dynamics and how it reorganizes with forecast horizon remain poorly understood.

Our findings show that incorporating subtype-resolved surveillance reveals a previously underappreciated structure in the predictability of seasonal influenza. While aggregate ILI+ signals contain strong short-term autoregressive information, they obscure heterogeneity in circulating viruses that becomes increasingly relevant at longer forecast horizons. By incorporating subtype information, we recover predictive structure that is not accessible from aggregated incidence alone. Importantly, predictability reorganizes across forecast horizons: short-term forecasts reflect substantial contributions from autoregressive persistence in recent incidence, whereas longer-horizon predictions increasingly depend on subtype-derived structure. Within this subtype component, the contribution of interaction-related features among co-circulating subtypes grows systematically with forecast horizon. These results suggest that a portion of the apparent unpredictability of influenza dynamics arises from aggregation of heterogeneous subtype processes, rather than from aggregate incidence trends alone. We further demonstrate through controlled simulation experiments that these patterns emerge in heterogeneous multi-subtype epidemic systems.

It is important to clarify that our objective was not to identify an optimal forecasting architecture. While more sophisticated models or ensemble strategies could be explored, the central aim of this study was to evaluate and decompose the informational value of subtype-resolved surveillance signals. We therefore selected a set of widely used and conceptually distinct data-driven model classes to ensure that the observed patterns reflect differences in informational content rather than model-specific optimization. The largely consistent results across model classes suggest that the contribution of subtype information is structural rather than algorithm-dependent.

The utility of subtype resolution aligns with established biological differences among influenza viruses and their co-circulation patterns. Influenza A(H3), A(2009 H1N1), and B differ in antigenic evolution, immunity landscapes, and age-specific attack patterns [34–36]. Recent work has shown that influenza subtype composition is itself structured and predictable [37], further supporting the idea that subtype-resolved signals may provide additional information for forecasting overall influenza activity. By combining epidemic intensity with subtype dynamics, the model captures how biologically distinct circulating viruses may contribute differently to epidemic growth without imposing explicit mechanistic assumptions. When multiple subtypes co-circulate, epidemic dynamics are shaped not only by their individual prevalence but also by their interaction within a shared host population [14, 38]. The increasing contribution of interaction-related features in our analysis is consistent with the idea that co-circulation structure may encode forward-looking information about aggregate epidemic trajectories. More broadly, our findings highlight the perspective that forecasting is not only a predictive task, but also a framework for understanding how predictive information is organized across timescales in complex epidemic systems. From a policy standpoint, these results support expanding subtype-resolved surveillance to improve epidemic preparedness and decision-making in increasingly data-driven public health systems.

Several limitations merit consideration. First, the effective training sample for subtype-informed forecasting was constrained by structural discontinuities in surveillance reporting. Changes in subtype categorization beginning in the 2015–2016 season, together with the exclusion of the COVID-19 disruption period, reduced the length of temporally consistent data. Nevertheless, the persistence of subtype-related contributions across periods, together with results from our simulation study, supports their structural relevance. Second, subtype resolution remains coarse relative to the genetic and antigenic diversity within circulating viruses; incorporating clade-level genomic information may further improve predictive performance [39]. Third, our analysis focuses on United States regional surveillance, and the extent to which these findings generalize to other climatic and surveillance contexts warrants further investigation. Finally, integrating information from co-circulating respiratory pathogens such as RSV or SARS-CoV-2 may provide a more comprehensive representation of epidemic interference dynamics [40].

## 4 Methods

### 4.1 Data sources and preprocessing

Weekly influenza surveillance data were obtained from the U.S. CDC FluView platform [11], covering all 10 U.S. HHS regions from the 2010–2011 through the 2024–2025 influenza seasons. ILI is defined by the CDC as fever (≥37.8^°^C) accompanied by cough or sore throat. In FluView, HHS-regional ILI percentages are reported as population-weighted aggregates based on state population. To approximate laboratory-confirmed influenza activity, we constructed the ILI+ metric as the product of the weighted ILI percentage and the proportion of respiratory specimens testing positive for influenza in clinical laboratories.

Virological data were harmonized into four stable categories: A(2009 H1N1), A(H3), A(other), and B. Reporting practices in FluView changed beginning in the 2015–2016 season, including the introduction of lineage-specific reporting for influenza B (BVic and BYam) and consolidation of influenza A subtype categories. To ensure longitudinal consistency, influenza B lineages were aggregated into a single B category. A(other) included unsubtyped influenza A specimens and rare variant strains. Subtype data were available at the HHS regional level. Although ILI was also reported at finer spatial resolutions (e.g., state level), we used the HHS-level ILI data to align with the spatial resolution of subtype reporting.

To avoid structural distortions associated with the COVID-19 pandemic, we excluded the period from epidemiological week 40 of 2018 through week 39 of 2022 from the primary analysis. During this interval, influenza circulation was substantially suppressed and surveillance practices were disrupted, limiting comparability with typical seasonal dynamics.

### 4.2 Descriptive summaries of subtype structure

Let *r* ∈ {1, …, 10} index the HHS regions and let *t* denote epidemiological week. For each subtype *k* ∈ { A(2009 H1N1), A(H3), A(other), B }, let Prop_*k,r,t*_ denote the proportion of laboratory-confirmed influenza-positive specimens in region *r* at week *t* that were attributed to subtype *k*.

We considered two complementary summaries of contemporaneous subtype structure (see also Fig. 2). First, we defined dominant subtype state based on the largest weekly subtype proportion. An observation was classified as *mixed* if no subtype accounted for more than 70% of proportional circulation; otherwise, it was assigned to the dominant subtype category corresponding to the subtype with the largest proportional circulation.

Second, to summarize overall co-circulation intensity, we defined the interaction measure

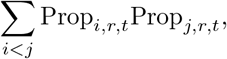

that is, the sum of all pairwise products of subtype proportions within a given region-week. Larger values indicate more balanced co-circulation across subtypes, whereas smaller values indicate stronger concentration in a single subtype. For the descriptive analyses in Fig. 2, observations were grouped according to tertiles of this co-circulation intensity measure, corresponding to low, medium, and high interaction.

For the descriptive analyses in Fig. 2, HHS region-week observations were grouped according to dominant subtype state and the corresponding interaction categories defined above. All subsequent trajectory summaries were computed within influenza seasons, so that ILI+_*r,t*+*h*_ was always evaluated relative to observations from the same season. For each region *r*, week *t*, and horizon *h* ∈ { 1, 2, 3, 4 }, we defined

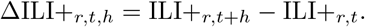

For each grouping category, Fig. 2 displays the mean of ΔILI+_*r,t,h*_ across observations at horizons *h* = 1, 2, 3, 4, together with 95% confidence intervals based on the standard error of the group mean.

### 4.3 Feature engineering

For forecasting analyses, the task was defined as predicting the out-of-sample ILI+ intensity in region *r* at week *t* + *h*, denoted by ILI+_*r,t*+*h*_, for horizons *h* = 1, 2, 3, 4.

Baseline models included only the most recent autoregressive observation, ILI+_*r,t*_. Subtype-informed models extended this feature set by incorporating contemporaneous subtype composition within the same HHS region. Specifically, we considered two types of subtype-derived predictors: subtype main-effect terms and subtype interaction terms.

Subtype main-effect terms were constructed as intensity-weighted subtype features of the form

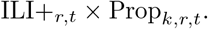

Subtype interaction terms were constructed as second-order features of the form

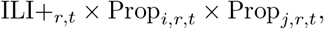

to account for potential co-circulation effects between subtypes.

### 4.4 Model frameworks and training

We implemented four predictive architectures to evaluate the incremental value of subtype-informed features: Random Forest (RF), Extreme Gradient Boosting (XGBoost), Long Short-Term Memory (LSTM), and Gated Recurrent Unit (GRU) networks.

Tree-based models (RF and XGBoost) were trained using standard regression objectives. Recurrent neural networks consisted of a single LSTM or GRU layer followed by a fully connected dense layer and a final output layer with a Rectified Linear Unit (ReLU) activation to enforce non-negative predictions. The number of hidden units was selected from {16, 24, 32 } via validation-based hyperparameter search. Models were optimized using RMSprop with kernel regularization. Early stopping with a patience of 10 epochs was applied based on validation RMSE.

Training and evaluation were conducted separately for pre- and post-COVID periods to account for structural changes in influenza dynamics. For the pre-COVID analysis, the training set covered epidemiological week 40 of 2010 through week 39 of 2015, the validation set spanned week 40 of 2015 through week 39 of 2017, and the testing set covered week 40 of 2017 through week 39 of 2018. For the post-COVID analysis, the training set included week 40 of 2010 through week 39 of 2018, the validation set covered week 40 of 2022 through week 39 of 2024, and the testing set spanned week 40 of 2024 through week 39 of 2025. For each forecast horizon, a separate model was trained and evaluated using the corresponding dataset split.

Models were trained jointly across all 10 HHS regions using pooled data, allowing parameter sharing across regions while preserving region-specific trajectories through the input features. Hyper-parameters were selected via random search minimizing validation RMSE. For tree-based models, the search space included the number of estimators, maximum tree depth, and subsampling rates. For recurrent neural networks, learning rates and batch sizes were tuned.

The ensemble forecast was constructed as the unweighted mean of the four individual model predictions, a simple and empirically robust aggregation strategy widely used in epidemic forecasting [41, 42].

### 4.5 Evaluation metrics

Forecasting performance was evaluated using four complementary metrics: RMSE, Pearson correlation coefficient, peak week error, and WIS.

RMSE measures the magnitude of prediction error:

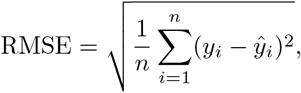

where *y*_*i*_ and *ŷ*_*i*_ denote the observed and predicted ILI+ values.

Pearson correlation quantifies the linear association between predicted and observed trajectories:

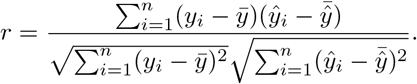

Peak Week Error measures the absolute difference between the predicted and observed epidemic peak week within each season:

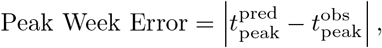

where 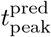 and 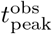 denote the predicted and observed peak week, respectively.

WIS evaluates the accuracy and calibration of probabilistic forecasts across multiple central prediction intervals. For a predictive distribution *F* with median *m* and observation *y*,

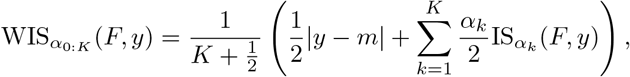

where the interval score is defined as

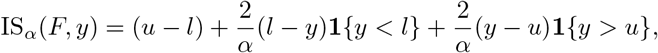

with [*l, u*] denoting the central (1 − *α*) prediction interval. We used *α* ∈ {0.02, 0.05, 0.10, 0.20, 0.30, 0.40, 0.50, 0.60, 0.70, 0.80, 0.90}, corresponding to central prediction intervals with nominal coverage 1 − *α*.

Predictive distributions were assumed Gaussian with mean *ŷ*_*t*_ and standard deviation *σ*_*t*_ [20, 43]. The standard deviation was estimated within each HHS region using a rolling window of the previous *k* = 10 out-of-sample residuals *e*_*t*_ = *y*_*t*_ − *ŷ*_*t*_:

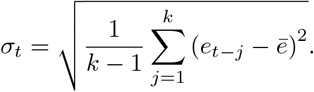

All metrics were first computed separately for each of the 10 HHS regions and then averaged across regions.

### 4.6 Feature importance analysis

Feature importance was quantified using permutation importance, defined as the increase in prediction error after randomly shuffling the values of a given feature while holding all other inputs fixed.

Formally, let ℒ (*ŷ, y*) denote the evaluation loss (RMSE in our case) computed on the testing set. For feature *X*_*j*_, we generate a perturbed dataset by randomly permuting its values across observations, denoted as 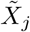, while leaving all other features unchanged. The permutation importance of feature *X*_*j*_ is defined as

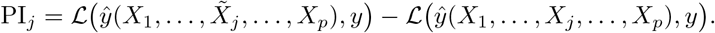

A larger PI_*j*_ indicates greater reliance of the model on feature *X*_*j*_ for predictive accuracy.

Importance scores were computed separately for each forecast horizon and for each of the four individual models. For the ensemble forecast, feature importance was summarized as the unweighted mean of the model-specific permutation importance scores across the four component models at the corresponding horizon.

To facilitate interpretation of predictive signal structure, features were grouped into three categories corresponding to the predictor construction described above: autoregressive incidence, subtype main-effect terms, and subtype interaction terms. Group-level importance was computed by summing permutation importance scores across all features within each category, and then normalizing by the total importance across all features for the corresponding model and forecast horizon.

## Data Availability

All data and code used in this analysis will be publicly available on a Github repository.

## Supplementary Information

Supplementary Information is available for this paper.

## Acknowledgements

M.S.Y.L., B.L., and K.K. were partially supported by cooperative agreement CDC-RFA-FT-23-0069 from the CDC’s Center for Forecasting and Outbreak Analytics. Its contents are solely the responsibility of the authors and do not necessarily represent the official views of the Centers for Disease Control and Prevention.

## Data Availability

All data and code used in this analysis will be publicly available on GitHub upon publication.

## S1 Region-specific patterns of subtype-related improvement

**Figure 1.**
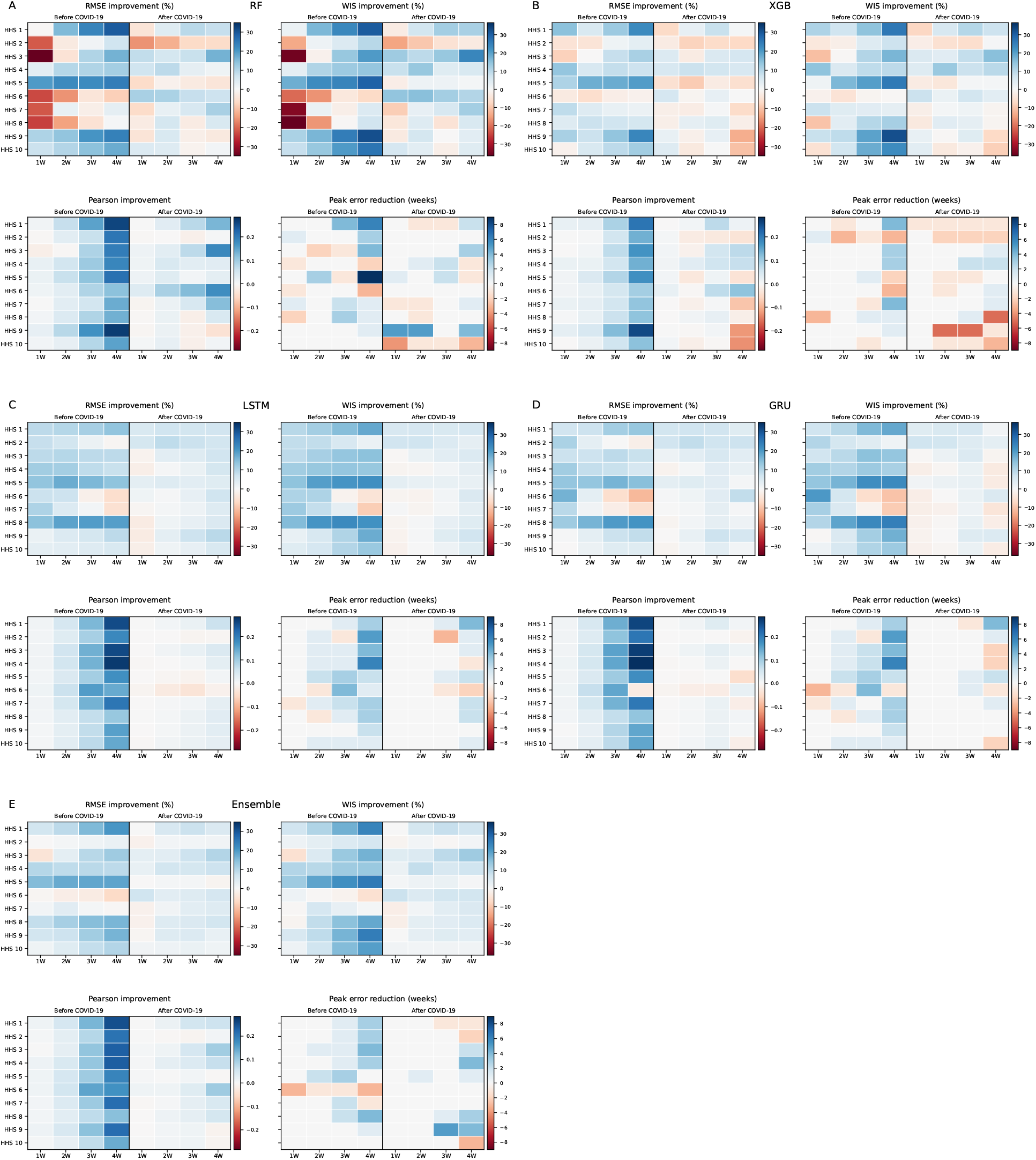
Heatmaps of region-specific subtype-related improvement across forecast horizons and model classes. Panels **A**–**E** correspond to RF, XGB, LSTM, GRU, and the ensemble model. Rows denote HHS regions and columns denote 1–4 week ahead horizons, shown separately for the pre-COVID and post-COVID periods. Color intensity indicates the magnitude of subtype-related improvement for RMSE, WIS, Pearson correlation, and peak error.

## S2 Extended-horizon forecasting results (5–8 weeks)

**Table 1.** Forecasting performance at 5–8 week ahead horizons. Results are shown separately for the pre-COVID and post-COVID periods. For RMSE and WIS, Δ% = 100 × (Subtype − Baseline)*/*Baseline. For Pearson Correlation and Peak Error, Δ = Subtype − Baseline. Negative Δ% indicates reduced error (RMSE/WIS) under the subtype model. Positive Δ in Pearson Correlation indicates improved correlation. Negative Δ in Peak Error indicates reduced peak timing error.

| Horizon | Model | RMSE |  |  | Pearson Correlation |  |  | Peak Error (weeks) |  |  | WIS |  |  |
| --- | --- | --- | --- | --- | --- | --- | --- | --- | --- | --- | --- | --- | --- |
| | | Baseline | Subtype | $\Delta\%$ | Baseline | Subtype | $\Delta$ | Baseline | Subtype | $\Delta$ | Baseline | Subtype | $\Delta\%$ |
| **Before COVID-19** |  |  |  |  |  |  |  |  |  |  |  |  |  |
| 5W | RF | 0.548 | 0.489 | -10.83 | 0.548 | 0.792 | 0.244 | 4.1 | 2.0 | -2.1 | 0.247 | 0.206 | -16.45 |
| 5W | XGB | 0.551 | 0.498 | -9.61 | 0.571 | 0.802 | 0.231 | 3.0 | 2.3 | -0.7 | 0.250 | 0.211 | -15.61 |
| 5W | LSTM | 0.571 | 0.562 | -1.44 | 0.550 | 0.809 | 0.259 | 4.2 | 1.5 | -2.7 | 0.250 | 0.236 | -5.91 |
| 5W | GRU | 0.570 | 0.566 | -0.63 | 0.547 | 0.781 | 0.235 | 4.5 | 1.7 | -2.8 | 0.250 | 0.237 | -5.10 |
| 5W | Ensemble | 0.557 | 0.522 | -6.43 | 0.561 | 0.840 | 0.279 | 4.8 | 1.7 | -3.1 | 0.247 | 0.217 | -12.36 |
| 6W | RF | 0.579 | 0.511 | -11.70 | 0.476 | 0.785 | 0.308 | 6.0 | 1.3 | -4.7 | 0.266 | 0.217 | -18.30 |
| 6W | XGB | 0.578 | 0.522 | -9.72 | 0.508 | 0.784 | 0.276 | 2.1 | 1.7 | -0.4 | 0.267 | 0.222 | -16.95 |
| 6W | LSTM | 0.606 | 0.593 | -2.25 | 0.450 | 0.744 | 0.294 | 6.5 | 1.7 | -4.8 | 0.268 | 0.251 | -6.51 |
| 6W | GRU | 0.606 | 0.600 | -1.03 | 0.445 | 0.728 | 0.283 | 6.2 | 2.0 | -4.2 | 0.269 | 0.253 | -5.85 |
| 6W | Ensemble | 0.590 | 0.550 | -6.78 | 0.484 | 0.827 | 0.343 | 6.1 | 2.2 | -3.9 | 0.264 | 0.230 | -13.11 |
| 7W | RF | 0.597 | 0.525 | -12.02 | 0.427 | 0.778 | 0.351 | 7.9 | 1.3 | -6.6 | 0.281 | 0.227 | -19.29 |
| 7W | XGB | 0.597 | 0.526 | -11.93 | 0.458 | 0.793 | 0.334 | 2.5 | 1.4 | -1.1 | 0.281 | 0.224 | -20.19 |
| 7W | LSTM | 0.630 | 0.612 | -2.93 | 0.333 | 0.652 | 0.318 | 6.7 | 2.1 | -4.6 | 0.285 | 0.265 | -6.85 |
| 7W | GRU | 0.634 | 0.617 | -2.55 | 0.326 | 0.662 | 0.335 | 6.5 | 2.1 | -4.4 | 0.286 | 0.265 | -7.51 |
| 7W | Ensemble | 0.611 | 0.564 | -7.75 | 0.420 | 0.818 | 0.399 | 6.9 | 1.8 | -5.1 | 0.279 | 0.239 | -14.40 |
| 8W | RF | 0.608 | 0.542 | -10.73 | 0.390 | 0.773 | 0.383 | 7.6 | 1.3 | -6.3 | 0.291 | 0.238 | -18.31 |
| 8W | XGB | 0.609 | 0.539 | -11.52 | 0.419 | 0.791 | 0.372 | 9.6 | 1.3 | -8.3 | 0.292 | 0.237 | -18.90 |
| 8W | LSTM | 0.647 | 0.631 | -2.43 | 0.173 | 0.598 | 0.425 | 7.0 | 2.6 | -4.4 | 0.299 | 0.273 | -8.76 |
| 8W | GRU | 0.642 | 0.638 | -0.59 | 0.112 | 0.581 | 0.469 | 8.0 | 2.7 | -5.3 | 0.312 | 0.275 | -11.71 |
| 8W | Ensemble | 0.622 | 0.582 | -6.56 | 0.342 | 0.816 | 0.474 | 7.7 | 1.3 | -6.4 | 0.294 | 0.250 | -15.01 |
| **After COVID-19** |  |  |  |  |  |  |  |  |  |  |  |  |  |
| 5W | RF | 0.559 | 0.545 | -2.49 | 0.553 | 0.599 | 0.045 | 4.7 | 4.6 | -0.1 | 0.252 | 0.235 | -7.01 |
| 5W | XGB | 0.559 | 0.570 | 2.05 | 0.556 | 0.514 | -0.042 | 4.2 | 4.0 | -0.2 | 0.254 | 0.245 | -3.54 |
| 5W | LSTM | 0.567 | 0.529 | -6.56 | 0.588 | 0.649 | 0.061 | 5.1 | 1.8 | -3.3 | 0.248 | 0.234 | -5.71 |
| 5W | GRU | 0.565 | 0.543 | -4.00 | 0.585 | 0.579 | -0.006 | 5.4 | 3.6 | -1.8 | 0.248 | 0.248 | 0.27 |
| 5W | Ensemble | 0.560 | 0.535 | -4.40 | 0.577 | 0.627 | 0.050 | 4.7 | 4.1 | -0.6 | 0.248 | 0.235 | -5.28 |
| 6W | RF | 0.592 | 0.564 | -4.64 | 0.475 | 0.547 | 0.072 | 5.4 | 6.4 | 1.0 | 0.275 | 0.250 | -9.06 |
| 6W | XGB | 0.593 | 0.576 | -2.80 | 0.480 | 0.504 | 0.024 | 3.2 | 4.9 | 1.7 | 0.279 | 0.254 | -8.66 |
| 6W | LSTM | 0.607 | 0.581 | -4.25 | 0.491 | 0.597 | 0.106 | 6.0 | 1.5 | -4.5 | 0.272 | 0.258 | -5.23 |
| 6W | GRU | 0.606 | 0.748 | 23.46 | 0.484 | -0.430 | -0.914 | 5.7 | 28.4 | 22.7 | 0.272 | 0.310 | 13.91 |
| 6W | Ensemble | 0.597 | 0.597 | -0.10 | 0.494 | 0.599 | 0.104 | 4.9 | 5.3 | 0.4 | 0.271 | 0.254 | -6.47 |
| 7W | RF | 0.637 | 0.582 | -8.69 | 0.314 | 0.510 | 0.196 | 8.3 | 5.0 | -3.3 | 0.311 | 0.267 | -14.21 |
| 7W | XGB | 0.617 | 0.583 | -5.40 | 0.410 | 0.510 | 0.099 | 3.5 | 5.5 | 2.0 | 0.300 | 0.268 | -10.70 |
| 7W | LSTM | 0.639 | 0.622 | -2.59 | 0.367 | 0.483 | 0.117 | 6.5 | 2.6 | -3.9 | 0.291 | 0.278 | -4.42 |
| 7W | GRU | 0.640 | 0.625 | -2.45 | 0.356 | 0.487 | 0.131 | 7.7 | 2.5 | -5.2 | 0.293 | 0.279 | -4.83 |
| 7W | Ensemble | 0.627 | 0.593 | -5.37 | 0.384 | 0.579 | 0.195 | 10.5 | 5.4 | -5.1 | 0.294 | 0.267 | -9.47 |
| 8W | RF | 0.633 | 0.594 | -6.26 | 0.307 | 0.524 | 0.217 | 16.2 | 6.1 | -10.1 | 0.315 | 0.281 | -10.51 |
| 8W | XGB | 0.630 | 0.604 | -4.10 | 0.358 | 0.478 | 0.120 | 6.0 | 6.9 | 0.9 | 0.313 | 0.286 | -8.69 |
| 8W | LSTM | 0.667 | 0.664 | -0.50 | 0.000 | 0.135 | 0.135 | 17.3 | 6.8 | -10.5 | 0.338 | 0.328 | -2.80 |
| 8W | GRU | 0.669 | 0.748 | 11.90 | -0.006 | -0.341 | -0.335 | 15.4 | 23.4 | 8.0 | 0.336 | 0.310 | -7.78 |
| 8W | Ensemble | 0.646 | 0.641 | -0.68 | 0.336 | 0.506 | 0.170 | 17.5 | 5.6 | -11.9 | 0.321 | 0.291 | -9.49 |

## S3 Supplementary robustness analysis: lagged subtype features

**Table 2.** Sensitivity analysis with subtype features lagged by one week relative to ILI+. Results are shown separately for the pre-COVID and post-COVID periods. For RMSE and WIS, Δ% = 100 × (Subtype − Baseline)*/*Baseline. For Pearson Correlation and Peak Error, Δ = Subtype − Baseline. Negative Δ% indicates reduced error (RMSE/WIS) under the subtype model. Positive Δ in Pearson Correlation indicates improved correlation. Negative Δ in Peak Error indicates reduced peak timing error.

| Horizon | Model | RMSE |  |  | Pearson Correlation |  |  | Peak Error (weeks) |  |  | WIS |  |  |
| --- | --- | --- | --- | --- | --- | --- | --- | --- | --- | --- | --- | --- | --- |
| | | Baseline | Subtype | $\Delta\%$ | Baseline | Subtype | $\Delta$ | Baseline | Subtype | $\Delta$ | Baseline | Subtype | $\Delta\%$ |
| **Before COVID-19** |  |  |  |  |  |  |  |  |  |  |  |  |  |
| 1W | RF | 0.220 | 0.238 | 8.17 | 0.948 | 0.954 | 0.006 | 1.3 | 1.1 | -0.2 | 0.090 | 0.098 | 8.57 |
| 1W | XGB | 0.231 | 0.226 | -2.05 | 0.942 | 0.953 | 0.011 | 1.2 | 1.1 | -0.1 | 0.097 | 0.097 | 0.01 |
| 1W | LSTM | 0.205 | 0.186 | -9.42 | 0.949 | 0.955 | 0.006 | 1.0 | 1.1 | 0.1 | 0.085 | 0.075 | -10.99 |
| 1W | GRU | 0.212 | 0.186 | -12.09 | 0.949 | 0.955 | 0.005 | 1.0 | 1.1 | 0.1 | 0.087 | 0.075 | -13.53 |
| 1W | Ensemble | 0.214 | 0.202 | -5.47 | 0.949 | 0.956 | 0.008 | 1.0 | 1.0 | 0.0 | 0.089 | 0.084 | -5.36 |
| 2W | RF | 0.354 | 0.344 | -2.84 | 0.841 | 0.884 | 0.042 | 2.1 | 1.7 | -0.4 | 0.147 | 0.142 | -3.83 |
| 2W | XGB | 0.355 | 0.349 | -1.66 | 0.845 | 0.877 | 0.031 | 1.6 | 1.7 | 0.1 | 0.151 | 0.148 | -2.37 |
| 2W | LSTM | 0.368 | 0.332 | -9.79 | 0.848 | 0.874 | 0.026 | 2.0 | 1.8 | -0.2 | 0.153 | 0.135 | -12.10 |
| 2W | GRU | 0.363 | 0.339 | -6.81 | 0.850 | 0.876 | 0.026 | 2.0 | 1.8 | -0.2 | 0.152 | 0.138 | -9.47 |
| 2W | Ensemble | 0.357 | 0.338 | -5.34 | 0.847 | 0.880 | 0.033 | 2.0 | 1.8 | -0.2 | 0.150 | 0.139 | -6.92 |
| 3W | RF | 0.447 | 0.420 | -5.96 | 0.736 | 0.823 | 0.086 | 3.0 | 2.2 | -0.8 | 0.190 | 0.175 | -8.33 |
| 3W | XGB | 0.448 | 0.423 | -5.70 | 0.745 | 0.813 | 0.068 | 2.2 | 1.7 | -0.5 | 0.196 | 0.178 | -8.88 |
| 3W | LSTM | 0.460 | 0.425 | -7.73 | 0.745 | 0.812 | 0.067 | 3.1 | 2.3 | -0.8 | 0.195 | 0.173 | -11.20 |
| 3W | GRU | 0.459 | 0.430 | -6.37 | 0.745 | 0.810 | 0.065 | 3.1 | 2.3 | -0.8 | 0.195 | 0.176 | -9.51 |
| 3W | Ensemble | 0.452 | 0.422 | -6.69 | 0.744 | 0.819 | 0.075 | 2.7 | 1.9 | -0.8 | 0.193 | 0.174 | -9.56 |
| 4W | RF | 0.507 | 0.468 | -7.65 | 0.624 | 0.782 | 0.158 | 3.7 | 1.8 | -1.9 | 0.221 | 0.195 | -12.04 |
| 4W | XGB | 0.512 | 0.468 | -8.62 | 0.648 | 0.779 | 0.131 | 2.6 | 1.9 | -0.7 | 0.227 | 0.197 | -13.35 |
| 4W | LSTM | 0.523 | 0.488 | -6.70 | 0.646 | 0.795 | 0.148 | 4.5 | 2.0 | -2.5 | 0.226 | 0.201 | -11.03 |
| 4W | GRU | 0.522 | 0.494 | -5.49 | 0.645 | 0.773 | 0.128 | 4.5 | 2.1 | -2.4 | 0.226 | 0.205 | -9.32 |
| 4W | Ensemble | 0.513 | 0.476 | -7.33 | 0.645 | 0.792 | 0.147 | 2.9 | 2.1 | -0.8 | 0.223 | 0.197 | -11.56 |
| **After COVID-19** |  |  |  |  |  |  |  |  |  |  |  |  |  |
| 1W | RF | 0.239 | 0.235 | -1.65 | 0.930 | 0.940 | 0.010 | 1.3 | 1.0 | -0.3 | 0.096 | 0.093 | -3.21 |
| 1W | XGB | 0.234 | 0.241 | 3.18 | 0.934 | 0.935 | 0.002 | 1.2 | 1.0 | -0.2 | 0.096 | 0.096 | -0.78 |
| 1W | LSTM | 0.232 | 0.226 | -2.46 | 0.939 | 0.938 | -0.001 | 1.0 | 1.0 | 0.0 | 0.093 | 0.091 | -2.33 |
| 1W | GRU | 0.230 | 0.223 | -2.96 | 0.939 | 0.939 | 0.000 | 1.0 | 1.0 | 0.0 | 0.092 | 0.092 | -0.91 |
| 1W | Ensemble | 0.231 | 0.226 | -2.10 | 0.937 | 0.940 | 0.004 | 1.0 | 1.0 | 0.0 | 0.093 | 0.090 | -2.95 |
| 2W | RF | 0.384 | 0.373 | -2.84 | 0.809 | 0.830 | 0.021 | 2.1 | 1.8 | -0.3 | 0.157 | 0.151 | -3.81 |
| 2W | XGB | 0.377 | 0.371 | -1.68 | 0.817 | 0.832 | 0.015 | 1.6 | 2.4 | 0.8 | 0.157 | 0.152 | -2.90 |
| 2W | LSTM | 0.377 | 0.366 | -2.89 | 0.827 | 0.826 | -0.001 | 2.0 | 2.0 | 0.0 | 0.153 | 0.151 | -1.19 |
| 2W | GRU | 0.376 | 0.366 | -2.69 | 0.826 | 0.826 | 0.000 | 2.0 | 2.0 | 0.0 | 0.153 | 0.151 | -0.99 |
| 2W | Ensemble | 0.376 | 0.364 | -3.37 | 0.821 | 0.832 | 0.011 | 2.0 | 1.9 | -0.1 | 0.154 | 0.149 | -3.05 |
| 3W | RF | 0.467 | 0.448 | -4.13 | 0.708 | 0.751 | 0.043 | 3.0 | 3.2 | 0.2 | 0.198 | 0.186 | -5.84 |
| 3W | XGB | 0.463 | 0.440 | -4.97 | 0.712 | 0.770 | 0.059 | 2.2 | 2.8 | 0.6 | 0.198 | 0.185 | -6.44 |
| 3W | LSTM | 0.463 | 0.447 | -3.48 | 0.731 | 0.731 | 0.000 | 3.1 | 2.9 | -0.2 | 0.193 | 0.189 | -2.04 |
| 3W | GRU | 0.462 | 0.446 | -3.52 | 0.730 | 0.735 | 0.005 | 3.1 | 2.9 | -0.2 | 0.193 | 0.188 | -2.37 |
| 3W | Ensemble | 0.461 | 0.439 | -4.78 | 0.723 | 0.754 | 0.032 | 2.7 | 2.9 | 0.2 | 0.194 | 0.184 | -5.07 |
| 4W | RF | 0.518 | 0.499 | -3.78 | 0.633 | 0.667 | 0.034 | 3.7 | 3.9 | 0.2 | 0.227 | 0.216 | -5.02 |
| 4W | XGB | 0.518 | 0.516 | -0.26 | 0.630 | 0.640 | 0.011 | 2.6 | 3.7 | 1.1 | 0.229 | 0.220 | -3.75 |
| 4W | LSTM | 0.520 | 0.496 | -4.66 | 0.660 | 0.669 | 0.009 | 4.5 | 3.3 | -1.2 | 0.222 | 0.216 | -3.10 |
| 4W | GRU | 0.518 | 0.498 | -3.91 | 0.658 | 0.671 | 0.013 | 4.5 | 3.5 | -1.0 | 0.222 | 0.215 | -3.32 |
| 4W | Ensemble | 0.516 | 0.494 | -4.20 | 0.649 | 0.684 | 0.035 | 2.9 | 3.2 | 0.3 | 0.223 | 0.213 | -4.70 |

## S4 Supplementary Methods: Simulation study

### S4.1 Epidemiological model

We conducted an additional simulation study to assess whether heterogeneous subtype dynamics can generate forecast-relevant co-circulation structure. We represented multi-subtype influenza circulation using three stylized co-circulating subtype components, indexed by *k* ∈ {1, 2, 3}. The parameter values for subtype 1, subtype 2, and subtype 3 were selected from, or calibrated to be consistent with, published influenza estimates so that they broadly approximate plausible A(H1N1)-, A(H3N2)-, and B-type seasonal influenza profiles, respectively.

For subtype *k* in region *r*, the dynamics were governed by a subtype-specific SEIR model:

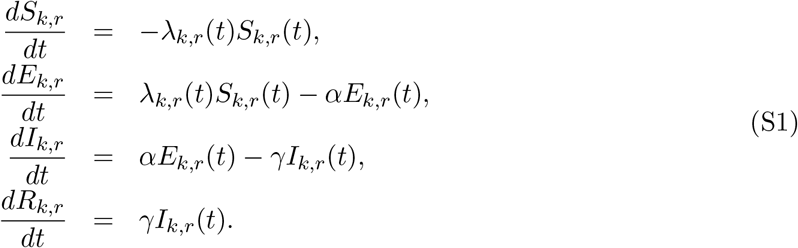

Here *S*_*k,r*_(*t*), *E*_*k,r*_(*t*), *I*_*k,r*_(*t*), and *R*_*k,r*_(*t*) denote the susceptible, exposed, infectious, and removed compartments for subtype *k* in region *r* at time *t*. The force of infection was defined as

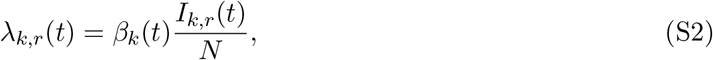

where *N* is the population size used as the scaling denominator. The transmission rate incorporated subtype-specific seasonal forcing:

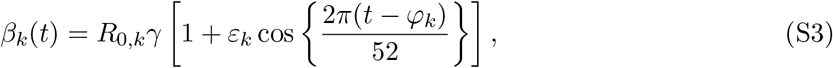

where *R*_0,*k*_ is the subtype-specific basic reproduction number, *γ* is the recovery rate, *ε*_*k*_ is the seasonal forcing amplitude, and *φ*_*k*_ is the subtype-specific seasonal phase offset.

The three subtype-specific systems used the same population size as the scaling denominator but evolved independently. Thus, *S*_*k,r*_(*t*) represents subtype-specific effective susceptibility rather than a single mutually exclusive susceptible compartment shared across all subtypes. Infection with one subtype did not deplete susceptibility to the other subtypes. This formulation allowed heterogeneous subtype-specific trajectories to arise from differences in transmissibility, timing, seasonality, initial susceptibility, and inter-seasonal perturbations.

### S4.2 Simulation parameters

The simulation parameters were chosen to generate stylized but epidemiologically plausible multi-subtype influenza dynamics. Parameters governing clinical time scales were based on published influenza estimates, whereas subtype-specific seasonal forcing amplitudes, phase offsets, regional structure, inter-seasonal perturbations, and observation noise were specified as simulation-design choices. Table 3 lists the parameter values and sources or modelling rationales.

For each simulated season and region, initial subtype-specific susceptibility fractions were sampled independently from Uniform(0.55, 0.80). Subtype-specific *R*_0_ values were perturbed by a season-level multiplicative factor sampled uniformly within *±*15% of the baseline subtype-specific value. This introduced inter-seasonal variability while preserving the intended subtype ranking in epidemic potential. Subtype 2 was assigned the highest baseline *R*_0_ to represent a subtype component with stronger epidemic potential, motivated by evidence that A(H3N2) exhibits relatively rapid antigenic drift and distinctive global circulation dynamics [6, 7].

**Table 3.** Simulation parameters and sources. Clinical time-scale parameters were chosen from published influenza estimates. Subtype 1, subtype 2, and subtype 3 were parameterized to broadly approximate plausible A(H1N1)-, A(H3N2)-, and B-type seasonal influenza profiles, respectively. Parameters marked as assumed were specified as stylized simulation-design choices to generate heterogeneous and asynchronous subtype-specific influenza dynamics rather than estimated from a specific historical outbreak.

| Parameter | Symbol | Value | Source / rationale |
| --- | --- | --- | --- |
| Population | $N$ | $10^6$ | Simulation-design choice |
| Latent period | $1/\alpha$ | 1.5 d; $\alpha = 4.67/\text{wk}$ | Approximate influenza incubation/latent timescale [1] |
| Infectious period | $1/\gamma$ | 3.0 d; $\gamma = 2.33/\text{wk}$ | Influenza shedding/transmission timescale [2, 3] |
| *Basic reproduction number* |  |  |  |
| Subtype 1 | $R_{0,1}$ | 1.8 | Chosen to broadly approximate an A(H1N1)-type seasonal influenza profile [4, 5] |
| Subtype 2 | $R_{0,2}$ | 2.2 | Chosen to broadly approximate a higher epidemic-potential A(H3N2)-type seasonal influenza profile [4, 6, 7] |
| Subtype 3 | $R_{0,3}$ | 1.6 | Chosen to broadly approximate an influenza B-type seasonal profile [4, 5] |
| *Seasonal forcing amplitude* |  |  |  |
| Subtype 1 | $\varepsilon_1$ | 0.20 | Assumed subtype-specific seasonal forcing |
| Subtype 2 | $\varepsilon_2$ | 0.25 | Assumed subtype-specific seasonal forcing |
| Subtype 3 | $\varepsilon_3$ | 0.15 | Assumed subtype-specific seasonal forcing |
| *Seasonal phase offset* |  |  |  |
| Subtype 1 | $\varphi_1$ | 0 weeks | Reference phase |
| Subtype 2 | $\varphi_2$ | +2 weeks | Assumed asynchronous subtype timing |
| Subtype 3 | $\varphi_3$ | −3 weeks | Assumed asynchronous subtype timing |
| Inter-annual $R_0$ perturbation | – | $\pm 15\%$ Uniform | Assumed inter-seasonal variability |
| *Simulation structure* |  |  |  |
| Regions | – | 5 | Simulation-design choice |
| Seasons | – | 30 | Simulation-design choice |
| Initial susceptibility | $S_k(0)/N$ | Uniform(0.55, 0.80) | Assumed range motivated by multi-strain influenza dynamics [8] |

### S4.3 Observation model and simulated outcomes

Weekly subtype-specific incidence was approximated by the latent-to-infectious transition flow,

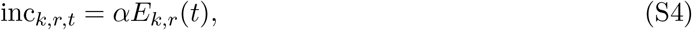

evaluated at weekly time points. To mimic measurement variability in surveillance data, subtype-specific Gaussian observation noise was added:

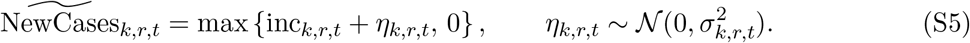

The noise standard deviation was defined as

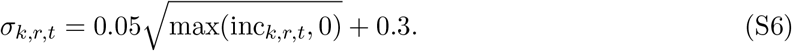

The aggregate simulated incidence was computed as

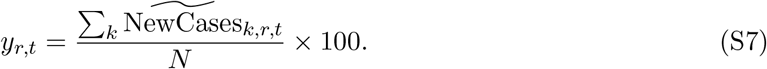

Subtype proportions were computed as

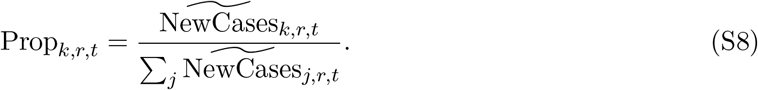

When the total simulated incidence was zero, subtype proportions were set to zero for all subtypes. The aggregate outcome *y*_*r,t*_ is analogous to the empirical ILI+ outcome in the forecasting pipeline, but it represents simulated infection incidence expressed as a percentage of the population rather than a syndromic surveillance measure.

## References

[1] Lafond, K.E., Nair, H., Rasooly, M.H., Valente, F., Booy, R., Rahman, M., Kitsutani, P., Yu, H., Guzman, G., Coulibaly, D., Armero, J., Jima, D., Howie, S.R.C., Ampofo, W., Mena, R., Chadha, M., Sampurno, O.D., Emukule, G.O., Nurmatov, Z., Corwin, A., Heraud, J.M., Noyola, D.E., Cojocaru, R., Nymadawa, P., Barakat, A., Adedeji, A., Horoch, M., Olveda, R., Nyatanyi, T., Venter, M., Mmbaga, V., Chittaganpitch, M., Nguyen, T.H., Theo, A., Whaley, M., Azziz-Baumgartner, E., Bresee, J., Campbell, H., Widdowson, M.-A., Group, G.R.H.P.P.G.W.: Global role and burden of influenza in pediatric respiratory hospitalizations, 1982–2012: A systematic analysis. PLOS Medicine 13(3), 1–19 (2016) 10.1371/journal.pmed.1001977

[2] Iuliano, A.D., Roguski, K.M., Chang, H.H., Muscatello, D.J., Palekar, R., Tempia, S., Cohen, C., Gran, J.M., Schanzer, D., Cowling, B.J., Wu, P., Kyncl, J., Ang, L.W., Park, M., Redlberger-Fritz, M., Yu, H., Espenhain, L., Krishnan, A., Emukule, G., Asten, L., Pereira da Silva, S., Aungkulanon, S., Buchholz, U., Widdowson, M.-A., Bresee, J.S., Global Seasonal Influenza-associated Mortality Collaborator Network: Estimates of global seasonal influenza-associated respiratory mortality: a modelling study. The Lancet 391(10127), 1285–1300 (2018) 10.1016/S0140-6736(17)33293-2

[3] Lipsitch, M., Finelli, L., Heffernan, R.T., and, G.M.L.: Improving the evidence base for decision making during a pandemic: The example of 2009 influenza a/h1n1. Biosecurity and Bioterrorism: Biodefense Strategy, Practice, and Science 9(2), 89–115 (2011) 10.1089/bsp.2011.0007

[4] Gandon, S., Day, T., Metcalf, C.J.E., Grenfell, B.T.: Forecasting epidemiological and evolutionary dynamics of infectious diseases. Trends in Ecology & Evolution 31(10), 776–788 (2016) 10.1016/j.tree.2016.07.010

[5] Viboud, C., Vespignani, A.: The future of influenza forecasts. Proceedings of the National Academy of Sciences 116(8), 2802–2804 (2019) 10.1073/pnas.1822167116

[6] Mathis, S.M., Webber, A.E., León, T.M., Murray, E.L., Sun, M., White, L.A., Brooks, L.C., Green, A., Hu, A.J., Rosenfeld, R., Shemetov, D., Tibshirani, R.J., McDonald, D.J., Kandula, S., Pei, S., Yaari, R., Yamana, T.K., Shaman, J., Agarwal, P., Balusu, S., Gururajan, G., Kamarthi, H., Prakash, B.A., Raman, R., Zhao, Z., Rodríguez, A., Meiyappan, A., Omar, S., Baccam, P., Gurung, H.L., Suchoski, B.T., Stage, S.A., Ajelli, M., Kummer, A.G., Litvinova, M., Ventura, P.C., Wadsworth, S., Niemi, J., Carcelen, E., Hill, A.L., Loo, S.L., McKee, C.D., Sato, K., Smith, C., Truelove, S., Jung, S.M., Lemaitre, J.C., Lessler, J., McAndrew, T., Ye, W., Bosse, N., Hlavacek, W.S., Lin, Y.T., Mallela, A., Gibson, G.C., Chen, Y., Lamm, S.M., Lee, J., Posner, R.G., Perofsky, A.C., Viboud, C., Clemente, L., Lu, F., Meyer, A.G., Santillana, M., Chinazzi, M., Davis, J.T., Mu, K., Pastore Y Piontti, A., Vespignani, A., Xiong, X., Ben-Nun, M., Riley, P., Turtle, J., Hulme-Lowe, C., Jessa, S., Nagraj, V.P., Turner, S.D., Williams, D., Basu, A., Drake, J.M., Fox, S.J., Suez, E., Cojocaru, M.G., Thommes, E.W., Cramer, E.Y., Gerding, A., Stark, A., Ray, E.L., Reich, N.G., Shandross, L., Wattanachit, N., Wang, Y., Zorn, M.W., Aawar, M.A., Srivastava, A., Meyers, L.A., Adiga, A., Hurt, B., Kaur, G., Lewis, B.L., Marathe, M., Venkatramanan, S., Butler, P., Farabow, A., Ramakrishnan, N., Muralidhar, N., Reed, C., Biggerstaff, M., Borchering, R.K.: Evaluation of FluSight influenza forecasting in the2021-22 and 2022-23 seasons with a new target laboratory-confirmed influenza hospitalizations. Nature Communications 15(1), 6289 (2024) 10.1038/s41467-024-50601-9

[7] Polonsky, J.A., Baidjoe, A., Kamvar, Z.N., Cori, A., Durski, K., Edmunds, W.J., Eggo, R.M., Funk, S., Kaiser, L., Keating, P., de Waroux, O.L.P., Marks, M., Moraga, P., Morgan, O., Nouvellet, P., Ratnayake, R., Roberts, C.H., Whitworth, J., Jombart, T.: Outbreak analytics: a developing data science for informing the response to emerging pathogens. Philosophical Transactions of the Royal Society B: Biological Sciences 374(1776), 20180276 (2019) 10.1098/rstb.2018.0276

[8] Biggerstaff, M., Alper, D., Dredze, M., Fox, S., Fung, I.C.-H., Hickmann, K.S., Lewis, B., Rosenfeld, R., Shaman, J., Tsou, M.-H., Velardi, P., Vespignani, A., Finelli, L., Influenza Forecasting Contest Working Group: Results from the Centers for Disease Control and Prevention’s Predict the 2013-2014 Influenza Season Challenge. BMC Infectious Diseases 16(1), 357 (2016) 10.1186/s12879-016-1669-x

[9] Reich, N.G., Brooks, L.C., Fox, S.J., Kandula, S., McGowan, C.J., Moore, E., Osthus, D., Ray, E.L., Tushar, A., Yamana, T.K., Biggerstaff, M., Johansson, M.A., Rosenfeld, R., Shaman, J.: A collaborative multiyear, multimodel assessment of seasonal influenza forecasting in the United States. Proceedings of the National Academy of Sciences 116(8), 3146–3154 (2019) 10.1073/pnas.1812594116

[10] Li, L., Yan, Z.L., Luo, L., Liu, W., Yang, Z., Shi, C., Ming, B.W., Yang, J., Cao, P., Ou, C.Q.: Influenza-associated excess mortality by age, sex, and subtype/lineage: Population-based time-series study with a distributed-lag nonlinear model. JMIR Public Health and Surveillance 9, 42530 (2023) 10.2196/42530

[11] Centers for Disease Control and Prevention: FluView: Influenza Surveillance Reports. https://www.cdc.gov/fluview/index.html. Accessed 11 February 2026 (2026)

[12] Bedford, T., Riley, S., Barr, I.G., Broor, S., Chadha, M., Cox, N.J., Daniels, R.S., Gunasekaran, C.P., Hurt, A.C., Kelso, A., Klimov, A., Lewis, N.S., Li, X., McCauley, J.W., Odagiri, T., Potdar, V., Rambaut, A., Shu, Y., Skepner, E., Smith, D.J., Suchard, M.A., Tashiro, M., Wang, D., Xu, X., Lemey, P., Russell, C.A.: Global circulation patterns of seasonal influenza viruses vary with antigenic drift. Nature 523(7559), 217–220 (2015) 10.1038/nature14460

[13] Petrova, V.N., Russell, C.A.: The evolution of seasonal influenza viruses. Nature Reviews Microbiology 16(1), 47–60 (2018) 10.1038/nrmicro.2017.118. Erratum in: Nat. Rev. Microbiol. 2018;16(1):60. doi: 10.1038/nrmicro.2017.146

[14] Ferguson, N.M., Galvani, A.P., Bush, R.M.: Ecological and immunological determinants of influenza evolution. Nature 422(6930), 428–433 (2003) 10.1038/nature01509

[15] Koelle, K., Cobey, S., Grenfell, B., Pascual, M.: Epochal evolution shapes the phylodynamics of interpandemic influenza A (H3N2) in humans. Science 314(5807), 1898–1903 (2006) 10.1126/science.1132745

[16] Latorre-Margalef, N., Brown, J.D., Fojtik, A., Poulson, R.L., Carter, D., Franca, M., Stallknecht, D.E.: Competition between influenza A virus subtypes through heterosubtypic immunity modulates re-infection and antibody dynamics in the mallard duck. PLoS Pathogens 13(6), 1006419 (2017) 10.1371/journal.ppat.1006419

[17] Kandula, S., Yang, W., Shaman, J.: Type- and subtype-specific influenza forecast. American Journal of Epidemiology 185(5), 395–402 (2017) 10.1093/aje/kww211

[18] Birrell, P.J., Zhang, X.-S., Corbella, A., van Leeuwen, E., Panagiotopoulos, N., Hoschler, K., Elliot, A.J., McGee, M., de Lusignan, S., Presanis, A.M., Baguelin, M., Zambon, M., Charlett, A., Pebody, R.G., De Angelis, D.: Forecasting the 2017/2018 seasonal influenza epidemic in England using multiple dynamic transmission models: a case study. BMC Public Health 20(1), 486 (2020) 10.1186/s12889-020-8455-9

[19] Turtle, J., Riley, P., Ben-Nun, M., Riley, S.: Accurate influenza forecasts using type-specific incidence data for small geographic units. PLoS Computational Biology 17(7), 1009230 (2021) 10.1371/journal.pcbi.1009230

[20] Tsang, T.K., Du, Q., Cowling, B.J., Viboud, C.: An adaptive weight ensemble approach to forecast influenza activity in an irregular seasonality context. Nature Communications 15(1), 8625 (2024) 10.1038/s41467-024-52504-1

[21] Centers for Disease Control and Prevention: FluSight Forecast Hub. https://github.com/cdcepi/FluSight-forecast-hub. Accessed 11 February 2026 (2026)

[22] Desai, A.N., Kraemer, M.U.G., Bhatia, S., Cori, A., Nouvellet, P., Herringer, M., Cohn, E.L., Carrion, M., Brownstein, J.S., Madoff, L.C., Lassmann, B.: Real-time epidemic forecasting: Challenges and opportunities. Health Security 17(4), 268–275 (2019) 10.1089/hs.2019.0022

[23] Breiman, L.: Random forests. Mach. Learn. 45(1), 5–32 (2001) 10.1023/A:1010933404324

[24] Chen, T., Guestrin, C.: Xgboost: A scalable tree boosting system. In: Proceedings of the 22nd ACM SIGKDD International Conference on Knowledge Discovery and Data Mining. KDD ‘16, pp. 785–794. Association for Computing Machinery, New York, NY, USA (2016). 10.1145/2939672.2939785

[25] Hochreiter, S., Schmidhuber, J.: Long short-term memory. Neural Computation 9(8), 1735–1780 (1997) 10.1162/neco.1997.9.8.1735

[26] Chung, J., Gulcehre, C., Cho, K., Bengio, Y.: Empirical Evaluation of Gated Recurrent Neural Networks on Sequence Modeling (2014). https://arxiv.org/abs/1412.3555

[27] Lessler, J., Reich, N.G., Brookmeyer, R., Perl, T.M., Nelson, K.E., Cummings, D.A.T.: Incubation periods of acute respiratory viral infections: a systematic review. The Lancet Infectious Diseases 9(5), 291–300 (2009) 10.1016/S1473-3099(09)70069-6

[28] Carrat, F., Vergu, E., Ferguson, N.M., Lemaitre, M., Cauchemez, S., Leach, S., Valleron, A.-J.: Time lines of infection and disease in human influenza: a review of volunteer challenge studies. American Journal of Epidemiology 167(7), 775–785 (2008) 10.1093/aje/kwm375

[29] Chan, L.Y.H., Morris, S.E., Stockwell, M.S., Bowman, N.M., Asturias, E.J., Rao, S., Lutrick, K., Ellingson, K.D., Nguyen, H.Q., Maldonado, Y., McLaren, S.H., Sano, E., Biddle, J.E., Smith-Jeffcoat, S.E., Biggerstaff, M., Rolfes, M.A.R., Talbot, H.K., Grijalva, C.G., Borchering, R.K., Mellis, A.M.: Estimating the generation time for influenza transmission using household data in the united states. Epidemics 50, 100815 (2025) 10.1016/j.epidem.2025.100815

[30] Truscott, J., Fraser, C., Cauchemez, S., Meeyai, A., Hinsley, W., Donnelly, C.A., Ghani, A., Ferguson, N.: Essential epidemiological mechanisms underpinning the transmission dynamics of seasonal influenza. Journal of the Royal Society Interface 9(67), 304–312 (2012) 10.1098/rsif.2011.0309

[31] Biggerstaff, M., Cauchemez, S., Reed, C., Gambhir, M., Finelli, L.: Estimates of the reproduction number for seasonal, pandemic, and zoonotic influenza: a systematic review of the literature. BMC Infectious Diseases 14, 480 (2014) 10.1186/1471-2334-14-480

[32] Yang, W., Lau, E.H.Y., Cowling, B.J.: Dynamic interactions of influenza viruses in hong kong during 1998–2018. PLoS Computational Biology 16(6), 1007989 (2020) 10.1371/journal.pcbi.1007989

[33] Perofsky, A.C., Huddleston, J., Hansen, C., Barnes, J.R., Rowe, T., Xu, X., Kondor, R., Went-worth, D.E., Lewis, N., Whittaker, L., Ermetal, B., Harvey, R., Galiano, M., Daniels, R.S., McCauley, J.W., Fujisaki, S., Nakamura, K., Kishida, N., Watanabe, S., Hasegawa, H., Sullivan, S.G., Barr, I.G., Subbarao, K., Krammer, F., Bedford, T., Viboud, C.: Antigenic drift and subtype interference shape A(H3N2) epidemic dynamics in the united states. eLife 13, 91849 (2024) 10.7554/eLife.91849.2

[34] Karageorgopoulos, D.E., Vouloumanou, E.K., Korbila, I.P., Kapaskelis, A., Falagas, M.E.: Age distribution of cases of 2009 (H1N1) pandemic influenza in comparison with seasonal influenza. PLoS One 6(7), 21690 (2011) 10.1371/journal.pone.0021690

[35] Gostic, K.M., Ambrose, M., Worobey, M., Lloyd-Smith, J.O.: Potent protection against H5N1 and H7N9 influenza via childhood hemagglutinin imprinting. Science 354(6313), 722–726 (2016) 10.1126/science.aag1322

[36] Arevalo, C.P., Le Sage, V., Bolton, M.J., Eilola, T., Jones, J.E., Kormuth, K.A., Nturibi, E., Balmaseda, A., Gordon, A., Lakdawala, S.S., Hensley, S.E.: Original antigenic sin priming of influenza virus hemagglutinin stalk antibodies. Proc. Natl. Acad. Sci. U. S. A. 117(29), 17221–17227 (2020) 10.1073/pnas.1920321117

[37] Bonacina, F., öBelle, P.-Y., Colizza, V., Lopez, O., Thomas, M., Poletto, C.: Characterization and forecast of global influenza subtype dynamics. Nature Health (2026) 10.1038/s44360-026-00069-2

[38] Kucharski, A.J., Andreasen, V., Gog, J.R.: Capturing the dynamics of pathogens with many strains. Journal of Mathematical Biology 72(1-2), 1–24 (2016) 10.1007/s00285-015-0873-4

[39] Luksza, M., Lässig, M.: A predictive fitness model for influenza. Nature 507(7490), 57–61 (2014) 10.1038/nature13087

[40] Nickbakhsh, S., Mair, C., Matthews, L., Reeve, R., Johnson, P.C.D., Thorburn, F., von Wissmann, B., Reynolds, A., McMenamin, J., Gunson, R.N., Murcia, P.R.: Virus-virus interactions impact the population dynamics of influenza and the common cold. Proc. Natl. Acad. Sci. U. S. A. 116(52), 27142–27150 (2019) 10.1073/pnas.1911083116

[41] Bleichrodt, A., Luo, R., Kirpich, A., Chowell, G.: Evaluating the forecasting performance of ensemble sub-epidemic frameworks and other time series models for the 2022-2023 mpox epidemic. R. Soc. Open Sci. 11(7), 240248 (2024) 10.1098/rsos.240248

[42] Amaral, A.V.R., Wolffram, D., Moraga, P., Bracher, J.: Post-processing and weighted combination of infectious disease nowcasts. PLoS Comput. Biol. 21(3), 1012836 (2025) 10.1371/journal.pcbi.1012836

[43] Aiken, E.L., Nguyen, A.T., Viboud, C., Santillana, M.: Toward the use of neural networks for influenza prediction at multiple spatial resolutions. Science Advances 7(25), 1237 (2021) 10.1126/sciadv.abb1237

## References

[1] Lessler, J., Reich, N. G., Brookmeyer, R., Perl, T. M., Nelson, K. E., and Cummings, D. A. T. (2009). Incubation periods of acute respiratory viral infections: a systematic review. The Lancet Infectious Diseases, 9(5), 291–300. 10.1016/S1473-3099(09)70069-6.

[2] Carrat, F., Vergu, E., Ferguson, N. M., Lemaitre, M., Cauchemez, S., Leach, S., and Valleron, A.-J. (2008). Time lines of infection and disease in human influenza: a review of volunteer challenge studies. American Journal of Epidemiology, 167(7), 775–785. 10.1093/aje/kwm375.

[3] Chan, L. Y. H., Morris, S. E., Stockwell, M. S., Bowman, N. M., Asturias, E. J., Rao, S., Lutrick, K., Ellingson, K. D., Nguyen, H. Q., Maldonado, Y., McLaren, S. H., Sano, E., Biddle, J. E., Smith-Jeffcoat, S. E., Biggerstaff, M., Rolfes, M. A. R., Talbot, H. K., Grijalva, C. G., Borchering, R. K., and Mellis, A. M. (2025). Estimating the generation time for influenza transmission using household data in the United States. Epidemics, 50, 100815. 10.1016/j.epidem.2025.100815.

[4] Truscott, J., Fraser, C., Cauchemez, S., Meeyai, A., Hinsley, W., Donnelly, C. A., Ghani, A., and Ferguson, N. (2012). Essential epidemiological mechanisms underpinning the transmission dynamics of seasonal influenza. Journal of the Royal Society Interface, 9(67), 304–312. 10.1098/rsif.2011.0309.

[5] Biggerstaff, M., Cauchemez, S., Reed, C., Gambhir, M., and Finelli, L. (2014). Estimates of the reproduction number for seasonal, pandemic, and zoonotic influenza: a systematic review of the literature. BMC Infectious Diseases, 14, 480. 10.1186/1471-2334-14-480.

[6] Bedford, T., Riley, S., Barr, I. G., Broor, S., Chadha, M., Cox, N. J., Daniels, R. S., Gunasekaran, C. P., Hurt, A. C., Kelso, A., Klimov, A., Lewis, N. S., Li, X., McCauley, J. W., Odagiri, T., Potdar, V., Rambaut, A., Shu, Y., Skepner, E., Smith, D. J., Suchard, M. A., Tashiro, M., Wang, D., Xu, X., Lemey, P., and Russell, C. A. (2015). Global circulation patterns of seasonal influenza viruses vary with antigenic drift. Nature, 523(7559), 217–220. 10.1038/nature14460.

[7] Petrova, V. N. and Russell, C. A. (2018). The evolution of seasonal influenza viruses. Nature Reviews Microbiology, 16(1), 47–60. 10.1038/nrmicro.2017.118.

[8] Yang, W., Lau, E. H. Y., and Cowling, B. J. (2020). Dynamic interactions of influenza viruses in Hong Kong during 1998–2018. PLoS Computational Biology, 16(6), e1007989. 10.1371/journal.pcbi.1007989.

